# Safety and Tolerability of Low Intensity Focused Ultrasound to the Anterior Insula in Patients with Fibromyalgia

**DOI:** 10.64898/2026.06.01.26354382

**Authors:** Aditya Kapoor, Yunruo Ni, Gabriel Isaac, David C. Keyes, Elizabeth A. Russo-Stringer, Wynn Legon

## Abstract

**Background:** Low-intensity focused ultrasound (LIFU) is an emerging noninvasive neuromodulation technique capable of targeting deep cortical and subcortical structures with high spatial precision. In healthy human volunteers, LIFU has demonstrated a favorable safety and tolerability profile across multiple studies. However, its safety and tolerability in clinical populations remains poorly characterized, representing a critical barrier to clinical translation. Here, we prospectively evaluate the safety and tolerability of LIFU targeting the left dorsal anterior insula (dAI) in patients with fibromyalgia (FM).

**Methods:** In a single-blind, sham-controlled, within-subjects crossover design, 13 individuals with FM (43.1 ± 13.2 years; 12 female) received 10 minutes of active LIFU (500 kHz, 1 kHz PRF, 36% duty cycle, 4.2 W/cm^2^ Isppa; 100 × 1-second pulse trains with a 5-second inter-train interval) targeting the left dorsal anterior insula (dAI) or sham on separate visits. Safety was evaluated through neuroradiological review of post vs. pre LIFU FLAIR MRI, quantitative voxel-wise FLAIR analysis, and patient report of symptoms (ROS). Tolerability was assessed using an experience assessment. Efficacy of the LIFU intervention was assessed using quantitative sensory testing (QST) including temporal summation of pain (TSP) and conditioned pain modulation (CPM).

**Results:** Neuroradiological review identified no new evidence of edema, microhemorrhage, acute ischemia, or white matter injury on post-LIFU structural imaging. Quantitative FLAIR analysis using contralateral-mirror-referenced relative FLAIR (rFLAIR) showed no significant within-subject change in the stimulated beam volume (ΔrFLAIR = 0.002 ± 0.025, t(12) = 0.30, P = 0.769, Cohen’s d_z_ = 0.08). No serious adverse events were documented and ROS indicated no change due to LIFU sonication. Participants rated the procedure as comfortable and could not distinguish active from sham LIFU. LIFU did not result in statistically significant changes for TSP (p = 0.797) or CPM (p = 0.465).

**Conclusions:** Ten minutes of LIFU targeting the left dAI was safe and well tolerated in individuals with FM, with no neuroradiological or quantitative MRI evidence of tissue effects and no serious adverse events. Blinding was preserved, and participants rated the procedure as comfortable. Although no significant changes were observed in experimental pain measures, these findings support the feasibility of targeting deep salience and pain amplification circuitry with LIFU in patients with FM and provide a foundation for adequately powered efficacy trials.

## INTRODUCTION

Low-intensity focused ultrasound (LIFU) is an emerging non-invasive neuromodulation technique that delivers mechanical energy through the skull into the brain, enabling reversible modulation of deep neural circuits with high spatial resolution [1,2]. This technique has been extensively characterized in small and large animal models [3–5], and a growing body of human literature now demonstrates that LIFU can safely and reversibly modulate human brain function [6–10]. Safety frameworks for LIFU for neuromodulation have evolved from diagnostic imaging, where decades of clinical use have established a strong safety record when exposures remain within established limits and are guided by risk–benefit principles [11]. Building on this foundation, the International Transcranial Ultrasonic Stimulation Safety and Standards (ITRUSST) consortium has recently proposed consensus thresholds for non-significant risk in human transcranial applications, including limits on mechanical index (MI ≤ 1.9) and conservative bounds on tissue heating (e.g., <2°C temperature rise and controlled thermal dose) [12]. These guidelines reflect current understanding that both cavitation-related mechanical effects and excessive heating represent the primary pathways for tissue injury, while appropriately controlled exposures are unlikely to produce structural damage.

Consistent with these principles, the safety profile of LIFU for neuromodulation in healthy adults is now relatively well characterized. Retrospective analyses report no serious adverse events [13] and systematic reviews of human transcranial ultrasound stimulation confirm a favorable safety and tolerability profile in healthy human volunteers [14,15]. However, despite increasing application of LIFU in clinical populations including mood, anxiety, and trauma-related disorders (MATRDs) [16,17], substance use disorder [18], and pain disorders [19], its safety and tolerability in patients with fibromyalgia (FM) remain unknown. Previous work in healthy volunteers has examined how LIFU applied to brain areas involved in pain processing, including the insula and anterior cingulate cortex, modulates pain-related responses [20–22] and recent studies suggest potential clinical benefit in pain populations [19]. These findings support the translational potential of LIFU for targeting circuits implicated in FM, yet prospective safety and tolerability data in this population are lacking. FM is a chronic pain disorder characterized by widespread musculoskeletal pain accompanied by fatigue, sleep disturbance, and cognitive difficulties [23]. Affecting approximately 2-6% of the population, FM imposes a substantial personal and societal burden, including reduced quality of life, increased healthcare utilization, and high rates of disability [24–27].

The insular cortex is a central node in the pathophysiology of fibromyalgia [28,29]. In the healthy brain, the insula integrates nociceptive [30], interoceptive [31], emotional [32], and salience-related signals [33], contributing to the subjective experience of pain [34]. In FM, the insula exhibits convergent structural, neurochemical, and functional abnormalities, including altered gray matter volume [35], excitatory–inhibitory imbalance [36,37] disrupted functional connectivity [28,38], and exaggerated responses to non-painful sensory input [39–42]. These alterations are particularly pronounced in the anterior insula (AI), where activity and connectivity are closely linked to clinical pain intensity and symptom severity [43,44], positioning the AI as a compelling target for neuromodulation.

Despite this strong mechanistic rationale, the safety and tolerability of AI-targeted LIFU in FM has not been established. As LIFU transitions toward clinical application, safety in patient populations cannot be inferred from healthy volunteer studies alone. For example, individuals with FM exhibit chronic alterations in cortical excitability and brain structure [35–37,39–41] that may influence responsiveness to neuromodulation. In clinical populations, identical NIBS protocols can produce blunted or directionally reversed responses relative to healthy controls [45,46]. In post-stroke pain patients specifically, NIBS-induced analgesia appears to operate by normalizing pathologically altered cortical excitability rather than inducing plasticity from a normal baseline as occurs in healthy controls [47]. Consistent with this, in FM, baseline neurochemistry predicts the magnitude of tDCS-induced response [48]. These findings indicate that NIBS efficacy in clinical pain populations is shaped by underlying disease state, motivating direct safety and tolerability evaluation in FM rather than inference from healthy-volunteer data. Additionally, clinical populations often present with comorbidities and medication use that are not represented in controlled experimental settings [49,50]. Although LIFU delivered within established biophysical thresholds is not expected to produce significant mechanical or thermal bioeffects [12] or serious adverse events, prospective safety data in chronic pain populations are lacking. Here, we report the first prospective, sham-controlled evaluation of the safety and tolerability of LIFU targeting the left dorsal anterior insula (dAI) in individuals with FM. Primary outcomes included neuroradiological assessment of pre- and post-LIFU Fluid-Attenuated Inversion Recovery (FLAIR) MRI, quantitative voxel-wise analysis of FLAIR signal [51–53], and participant-reported symptoms [13,54] and tolerability. In addition, we assessed changes in experimental pain processing, including temporal summation of pain (TSP) and conditioned pain modulation (CPM). TSP indexes the progressive amplification of pain perception during repeated nociceptive input and serves as a psychophysical readout of pain amplification, which is the heightened responsiveness of central nociceptive neurons that drives augmented pain processing in nociplastic conditions including FM [43,44,55]. CPM indexes the descending inhibitory control of pain, in which a noxious conditioning stimulus attenuates the perceived intensity of a subsequent test stimulus [56,57]. Both processes are dysfunctional in FM patients as they exhibit enhanced TSP at lower stimulus intensities than healthy controls [42,58] and impaired CPM, often showing facilitation of perceived intensity of the test stimulus rather than expected inhibition [59,60]. This study was designed to establish the safety and feasibility of AI-directed LIFU in FM and to provide a foundation for future efficacy trials.

## METHODS

### Participants

All experimental procedures were approved by the Virginia Tech Institutional Review Board (VT-IRB 22-1597), and the study was registered on ClinicalTrials.gov (NCT05751096). Nineteen participants were enrolled and completed baseline structural MR and CT imaging. One participant was excluded due to inability to complete the required structural MRI, and five were lost to follow-up due to scheduling conflicts or participant unavailability prior to receiving any intervention, yielding a final sample of 13 participants who completed the full experimental protocol (mean age = 43.1 ± 13.2 years; range = 24–65 years; 12 females, 1 male). All participants met inclusion and exclusion criteria, provided written informed consent, and received financial compensation for participation. Inclusion criteria required adults aged 18–65 years with a confirmed diagnosis of fibromyalgia established by a physician using the American College of Rheumatology 2011 modified fibromyalgia criteria. Participants were required to be English-speaking and capable of providing informed consent. Exclusion criteria were consistent with established contraindications to noninvasive neuromodulation [61] including contraindications to MRI or CT, history of neurological disorders (e.g., Parkinson’s disease, epilepsy), history of head injury with loss of consciousness >10 minutes, active medical conditions or treatments with potential central nervous system effects (e.g., Alzheimer’s disease), history of seizures or pseudo-seizures, use of medications that lower seizure threshold, diagnosis or history of Raynaud’s disease, peripheral neuropathy or diabetes mellitus, pregnancy, history of alcohol or drug dependence, and allergy to materials in the quantitative sensory testing device.

#### Baseline Clinical Assessments

To characterize baseline clinical symptoms, participants completed validated self-report assessments prior to any experimental procedures. Overall FM impact was assessed using the Revised Fibromyalgia Impact Questionnaire [62]. Physical function, sleep disturbance, and pain quality were assessed using short forms from the Patient-Reported Outcomes Measurement Information System (PROMIS) [63–65]. Physical function was measured using the PROMIS Physical Function 6-item short form, sleep disturbance using the PROMIS Sleep Disturbance Short Form 6a, and nociceptive and neuropathic pain qualities using the PROMIS Adult v2.0 Nociceptive and Neuropathic Pain Quality Short Form 5a instruments.

Pain-related cognitive and functional factors were further assessed using the Pain Catastrophizing Scale (PCS) [66]. Pain interference and functional impact were assessed using the PEG (Pain, Enjoyment, General Activity) [67]. Symptoms of anxiety and depression were screened using the 2-item Generalized Anxiety Disorder scale (GAD-2) and the 2-item Patient Health Questionnaire (PHQ-2), respectively [68,69]. Substance use risk was assessed using the Tobacco, Alcohol, Prescription Medication, and Other Substance Use (TAPS-2) Tool [70].

### Overall study design

The study used a sham-controlled, within-subjects crossover, pre/post design (see **Figure 1**). Participants completed four visits in total: one baseline visit, two intervention visits (one LIFU and one sham), and one dedicated safety imaging visit (see MRI and CT Imaging section).

**Figure 1.**
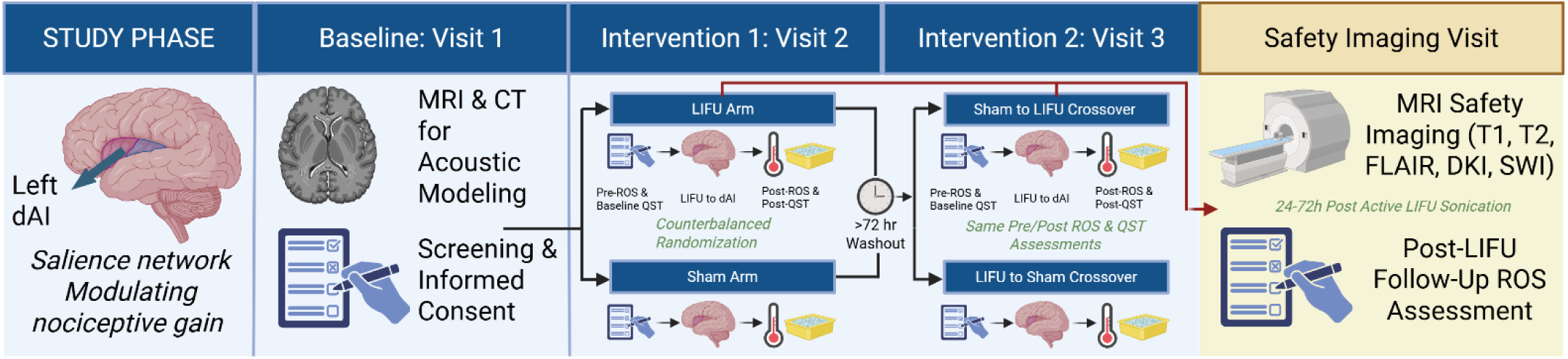
Study design and visit structure. Participants completed a baseline visit (Visit 1) that included informed consent and MRI and CT acquisition for screening and acoustic modeling. At Visit 2 and Visit 3, separated by a minimum 72-hour washout, participants received LIFU or sham sonication to the left dorsal anterior insula (dAI) in counterbalanced order. Each intervention visit included pre- and post-sonication (at least 30 minutes after sonication) administration of the Review of Systems (ROS) and quantitative sensory testing (QST). The red arrow indicates that the dedicated safety imaging visit, comprising T1, T2, FLAIR, DKI, and SWI sequences and a post-LIFU follow-up ROS assessment, was anchored to the active LIFU sonication visit and completed within 24–72 hours of sonication, regardless of intervention order.

During the baseline visit (Visit 1), participants provided written informed consent and completed baseline clinical assessments. Structural brain MRI and CT scanning were also acquired for individualized LIFU targeting, acoustic modeling (see Individual and Group Acoustic Modelling section), and baseline safety imaging (see Safety Imaging section).

The two formal intervention visits (Visits 2 and 3) each consisted of active LIFU or sham neuromodulation targeting the left dorsal anterior insula, delivered in counterbalanced order, such that approximately half received sham first and half received LIFU first. The dedicated safety imaging visit was always anchored after the active LIFU intervention and was scheduled 24–72 hours after active sonication, regardless of whether participants received LIFU or sham first. There was a minimum of 72 hours between the LIFU and sham visits to allow adequate washout of any neuromodulation carryover effects, as evidence suggests effects may persist for several hours [71], as well as provide enough time to complete the dedicated post-LIFU safety imaging visit. During each intervention visit, participants first completed a Report of Symptoms (ROS) assessment. Participants then completed baseline TSP and CPM testing, with each task lasting approximately 10 minutes, separated by 5-minute rest periods. Participants then received 10 minutes of active LIFU or sham neuromodulation. Approximately 2 minutes after sonication, TSP and CPM testing were conducted in counterbalanced order. At the end of each intervention visit, participants completed a post-ROS assessment at least 30 minutes after active or sham sonication. Each visit lasted approximately 3 hours. A follow-up ROS assessment was additionally administered at the safety imaging visit, 24-72 hours after LIFU, to further evaluate potential symptom changes following active LIFU sonication.

### MRI and CT Imaging

At the baseline visit, the full safety imaging battery (detailed below) was acquired to serve as the pre-sonication reference, along with a CT scan used for individualized acoustic modeling. CT scans were acquired with Kernel = Hr60, FoV = 250 mm, kV = 120, rotation time = 1 second, pitch = 0.55, caudocranial acquisition, 1.0 mm increments for a total of 121 images. All MRI data were acquired on a Siemens 3T Prisma scanner (Siemens Medical Solutions, Erlangen, Germany) at the Fralin Biomedical Research Institute’s Human Neuroimaging Laboratory.

#### Anatomical safety scans

A battery of anatomical scans that included a T1-weighted MPRAGE, T2-weighted, 3D FLAIR, DKI, and SWI (details below) were collected pre- and post- (24 – 72 hours) the LIFU intervention. These scans were taken to evaluate potential structural changes from the LIFU intervention including microhemorrhages, edema, acute ischemia, and white matter injury. The timing of the post scans was selected based on the timing or evolution of vasogenic and cytotoxic edema, the most likely subacute MRI-detectable tissue responses, that typically reach peak FLAIR signal intensity within 24–72 hours of an injurious event [72–75]. All pre and post images were reviewed and compared by a board-certified neuroradiologist (D.C.K). For participants assigned to LIFU-first (n=7), this dedicated safety imaging visit occurred before the sham intervention visit; for participants assigned to sham-first, it occurred after the LIFU intervention visit (see **Figure 1**).

The details of the safety imaging battery are: T1-weighted MPRAGE (TR = 2300 ms, TI = 900 ms, TE = 2.98 ms, flip angle = 9°, 1.0 mm slice thickness, matrix = 256 × 240), T2-weighted (TR = 3200 ms, TE = 565 ms, flip angle = 120°, 1.0 mm slice thickness, matrix = 256 × 240), 3D FLAIR (TR = 4800 ms, TI = 1650 ms, TE = 441 ms, flip angle = 120°, 1.0 mm slice thickness, matrix = 256 × 240), DKI (TR = 4100 ms, TE = 82 ms, flip angle = 90°, 1.7 mm slice thickness, b = [0, 1500, 3000]), and SWI (TR = 27 ms, TE = 20 ms, flip angle = 15°, 1.5 mm slice thickness, matrix = 256 × 232). No contrast agent was administered.

### LIFU Transducers and Waveform

Ultrasound waveforms were generated with a dual-channel function generator (BK4078B, Precision Instruments). Channel 1 produced a 5 V peak-to-peak, 1 kHz square-wave pulse with a 36% duty cycle, which was used to gate Channel 2. Channel 2 generated a 500 kHz sinusoidal waveform that was transmitted through a 100 W linear RF amplifier (E&I 2100L, Electronics & Innovation) prior to delivery to the ultrasound transducer. LIFU neuromodulation consisted of 100 pulse trains, each lasting 1 second, delivered with a fixed pulse train interval of 5 seconds, resulting in a total neuromodulation duration of 10 minutes. Given a 36% duty cycle during each 1-second sonication and the 5-second inter-stimulus interval, the resulting average duty cycle across the full neuromodulation period was 6%, totaling 36 seconds of active sonication. These parameters were selected based on our prior work demonstrating modulation of temporal summation of pain using this waveform applied to the dorsal anterior insula in healthy volunteers [22].

The applied free-field peak rarefactional pressure was 380 kPa, corresponding to a spatial-peak pulse-average intensity (Isppa) of 4.2 W/cm², a spatial-peak temporal-average intensity (Ispta) of 1.5 W/cm², and a mechanical index (MI) of 0.20.

Three different 500 kHz focused ultrasound transducers (Sonic Concepts, Bothell, WA) were used to accommodate individual variability in scalp-to-target depth. An H-281 (single-element, active diameter 45.0 mm, geometric focus 45.0 mm, focal depth from exit plane 38.0 mm) was used in nine participants. An H-104 (single-element, active diameter 64.0 mm, geometric focus 63.2 mm, focal depth from exit plane 52.0 mm) was used in three participants and a CTX-500 (dual-element annular array, active diameter 60.0 mm, geometric focus 64.0 mm, electronically adjustable focal depth) was used in one participant due to temporary unavailability of the H-281. The same ultrasound waveform and output pressure was used across all three transducers. Participant specific target depths are reported in the Results.

The ultrasound transducers were acoustically coupled to the participant’s scalp by first applying ultrasound gel to separate the hair, expose the skin surface, and reduce the presence of air at the interface. A custom-designed coupling puck was then placed between the transducer and the scalp to further provide acoustic coupling [76]. Pucks were fabricated with variable thicknesses, enabling precise adjustment of the acoustic focal depth to account for individual variability in target depth.

The left dorsal anterior insula target was defined for each participant using their individual structural MRI, guided by an insular atlas [77]. Target depth relative to the scalp was quantified using a neuronavigation system (BrainSight, Rogue Research, Montreal, QC, Canada). The target location corresponded to the left anterior short gyrus.

Transducer positioning on the scalp was guided and continuously monitored using the BrainSight neuronavigation system. LIFU neuromodulation was initiated only when the estimated scalp placement error was below 2 mm. Sham neuromodulation was delivered using the same transducer and coupling procedures as in the active visit, but the ultrasound beam was blocked by an acoustically opaque attenuator [78]. All other procedures were matched to the active LIFU condition, including neuronavigation, application of ultrasound gel and coupling puck, and presentation of continuous white noise for auditory masking (see Auditory Masking section for additional details).

#### Empirical Acoustic Field Mapping

Acoustic pressure fields were characterized in a degassed water tank using a calibrated needle hydrophone (HNR-0500, Onda Corp., Sunnyvale, CA) mounted on a motorized 3D positioning stage. Lateral (XY) and axial (YZ) planar scans were acquired at 0.25 mm resolution to define the focal spot geometry and full-width at half-maximum (FWHM) dimensions of each transducer. Input-output sweeps across the operating voltage range established the linear relationship between driving voltage and acoustic pressure at the focus for each. These measurements were used to calibrate the driving voltage for in vivo visits such that the applied extracranial pressure was standardized across all participants.

### Acoustic Modelling

Subject-specific computational acoustic models were constructed using individual structural MRI and CT data to characterize LIFU propagation through the skull and the resulting intracranial pressure fields. Simulations were performed in MATLAB using the k-Wave toolbox [79], which solves the full acoustic wave equation using a pseudospectral time-domain approach on a discretized spatial grid. CT images were used to derive skull acoustic properties, while T1-weighted MRI provided anatomical guidance for individualized targeting of the insular cortex based on each subject’s anatomy. CT volumes were registered to MRI space using FSL FLIRT [80]. The skull was segmented from CT images using intensity-based thresholding, and the intracranial volume was modeled as water, given the relatively small impedance mismatches among soft tissues at the operating frequency [81]. The co-registered CT and MRI volumes were then resampled at seven points per wavelength onto the k-Wave computational grid for acoustic simulation. Voxel-wise acoustic parameters within the skull were estimated from CT Hounsfield units assuming a linear relationship with skull porosity and density [82]. The ultrasound transducers were explicitly modeled to reproduce empirically measured free-field pressure distributions obtained from acoustic tank calibration, consistent with prior validation studies [83]. Acoustic modelling details have been described previously [84].

Following whole-head acoustic simulations, subject-specific pressure maps and the corresponding kgrid-space structural MRI volumes were exported in NIfTI format. To return simulation outputs to anatomical space, each kgrid-space MRI volume was registered to the participant’s native FreeSurfer-reconstructed T1 volume using a two-stage approach: an initial affine alignment followed by nonlinear refinement using ANTs symmetric normalization (SyN) [85]. The resulting warp fields were then applied to the simulated pressure maps using the same transformations to ensure spatial correspondence. Brain masks were generated from the FreeSurfer brainmask volume (eroded by 3 mm to exclude skull-edge voxels). Region-of-interest analyses focused on the left dorsal anterior insula (dAI), defined using the “left agranular insula” in the Brainnetome atlas [86], thresholded at 70% probability to generate a binary mask and warped to each participant’s T1 space via nonlinear registration. For each participant, the left dAI mask was applied to the subject-specific pressure map. To quantify per-participant spatial overlap between the LIFU beam and the dAI target and to characterize individualized intracranial exposure, we applied two pressure thresholds to each subject’s intracranial pressure map: −3 dB (∼71% of intracranial peak; the high-pressure focal core) and full width at half maximum (FWHM; 50% of intracranial peak). For each threshold, we computed ROI coverage (proportion of dAI voxels exceeding threshold), peak pressure within the ROI, and mean pressure within suprathreshold regions.

### Thermal Modelling

Thermal simulations were performed using the modified mixed-domain method (mSOUND) with a three-layer skull model (cortical-trabecular-cortical) based on Benchmark 6 from Aubry et al [87], solving for temperature fields from the simulated acoustic field via the bioheat equation using k-Wave diffusion [87]. The simulation was run at an extracranial input pressure of 1 MPa, corresponding to a conservative, higher-than-experimental exposure. For computational feasibility, the experimental 1 kHz PRF was compressed to 10 Hz PRF while holding total acoustic energy constant; prior work has demonstrated that lowering PRF in this manner produces negligible differences in predicted thermal rise [88]. The simulated pulse cycle was 36 ms on and 64 ms off, corresponding to a 10 Hz PRF and 36% duty cycle, delivered for 600 seconds total. To provide a conservative upper-bound estimate of heating, the simulation assumed 65% absorption through the skull with all absorbed energy converted to heat.

### Safety and Tolerability Assessments

Primary safety outcomes were: (i) neuroradiological review of pre- and post-LIFU structural MRI; (ii) quantitative voxel-wise analysis of pre- and post-LIFU FLAIR imaging; and (iii) symptoms queried via the Report of Symptoms (ROS). A serious adverse event (SAE) was defined as any untoward medical occurrence resulting in death, life-threatening illness, inpatient hospitalization or prolongation thereof, persistent or significant disability, or any event judged medically important enough to require intervention. Per the study protocol (VT-IRB 22-1597), pre-specified participant-level stopping criteria were (i) evidence of microhemorrhage, edema, acute ischemia, or white matter injury on post-LIFU structural imaging, (ii) a new severe symptom on the post-sonication ROS that was absent at pre-visit baseline and judged by PI review to be attributable to the intervention, (iii) a SAE. Symptoms on the ROS are characterized descriptively by severity (absent, mild, moderate, severe), consistent with prior LIFU safety reporting [13].

#### Neuroradiological Review

Pre- and post-LIFU structural MRI from each participant were reviewed by a board-certified neuroradiologist (D.C.K.) using the 3D T2 FLAIR source images). For each participant, the baseline (pre-LIFU) scan and the post-LIFU safety imaging scan, acquired 24–72 hours after sonication, were read together to identify any new findings on the post-LIFU scan relative to baseline. Findings of interest were defined per the pre-specified stopping criteria (microhemorrhage, edema, acute ischemia, or white matter injury) and were further evaluated for incidental abnormalities including masses, hemorrhage, extra-axial fluid collections, midline or other congenital/developmental abnormalities, hydrocephalus, ventricular entrapment, and herniation. Findings were categorized as new versus pre-existing and stable based on direct comparison of the pre- and post-LIFU sequences.

#### Quantitative FLAIR preprocessing

Paired Pre- and post-LIFU 3D FLAIR images were reoriented to standard orientation, and inferior slices below the cerebellum were removed, and inhomogeneities were corrected using N4BiasFieldCorrection (ANTs) [89]. Each bias-corrected FLAIR images was then co-registered to each participant’s FreeSurfer-reconstructed T1 space using FSL FLIRT with a rigid-body transform and the correlation-ratio cost function. Subject-specific whole-brain mask derived by FreeSurfer was eroded by 2 voxels to exclude edges prone to registration and partial-volume artifacts. Within each subject’s brain mask, both pre/post FLAIR volumes were intensity-normalized, yielding maps *S*_pre_(*x*)and *S*_post_(*x*), where *x* indexes voxels in the participant’s T1 space. These maps were used in all downstream analyses.

#### Quantitative FLAIR analysis

Localized signal changes at the left hemisphere dorsal anterior insula (lh-dAI) ROI were quantified using the relative FLAIR signal intensity (rFLAIR) framework [51,90]. In this framework, the FLAIR signal at each voxel is expressed as a ratio to the median FLAIR signal in a contralateral homologous reference region. The reference region *R* was the right-hemisphere dorsal anterior insula (rh-dAI), generated from the Brainnetome atlas using the same nonlinear-registration pipeline and 70% atlas probability threshold applied to the lh-dAI ROI mask. The target region *B* was the full-width at half-maximum (FWHM) intracranial beam volume, defined as voxels at which the simulated acoustic pressure field exceeded 50% of its intracranial maximum, intersected with the brain mask.

Per-voxel rFLAIR was computed independently for each session as:

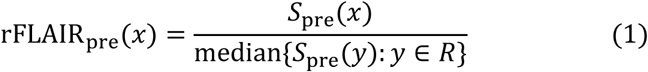

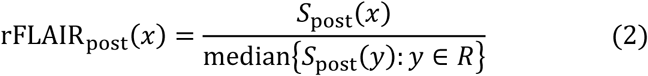

Voxel-wise change between visits was

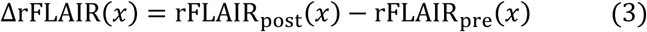

For per-subject group-level summaries, scalar pre- and post-LIFU rFLAIR within the FWHM beam were computed as the mean of the voxel-wise rFLAIR map across *B*, which is expressed as

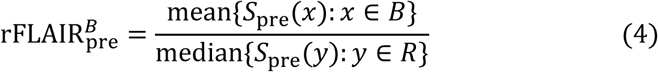

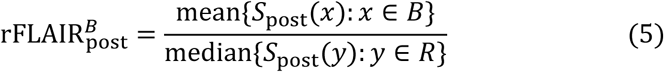

At the group level, Wilcoxon signed-rank test evaluated whether 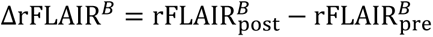 differed from zero. Paired Cohen’s *d*_*z*_was reported as the effect-size estimate, with 95% confidence intervals on the group-mean ΔrFLAIR^*B*^.

#### Report of Symptoms

A Report of Symptoms (ROS) assessment [12,54] queried the presence and severity of 17 symptoms on a 4-point ordinal scale (0 = absent, 1 = mild, 2 = moderate, 3 = severe). The term ‘severe’ as used on the ROS, denoted by ‘3’ on the 4-point scale, is independent of the aforementioned definition of a SAE and is meant to convey only the highest severity of the respective symptom.

At each intervention visit, the ROS was administered at the beginning of the study visit, before sonication (pre-sonication), and at least 30 minutes after sonication (post-sonication). A follow-up ROS to evaluate longitudinal safety of LIFU was administered at the dedicated safety imaging visit 24 – 72 hours following LIFU (see Overall Study Design and **Figure 1**).

For each symptom, a within-visit, LIFU or sham intervention, change score was calculated by subtracting the pre-sonication rating from the post-sonication rating, such that positive values indicated worsening and negative values indicated improvement. Symptoms are reported descriptively by severity (absent, mild, moderate, severe) across three timepoints — pre-sonication, post-sonication, and follow-up — consistent with prior LIFU safety reporting [13]. Follow-up symptom burden is reported as change from the active LIFU post-visit ratings to the follow-up assessment, to evaluate for any potential sustained effects of LIFU.

Two categories of new symptom elevations were enumerated separately for each condition. For within-visit assessments, new instances were defined relative to the pre-sonication baseline: (i) new severe instances, defined as a symptom rated absent at pre-sonication baseline that reached severe post-sonication, and (ii) new moderate instances, defined as a symptom rated absent at pre-sonication baseline that reached moderate post-sonication. New severe within-visit instances were reviewed against pre-specified stopping criteria, and any such elevation triggered review to assess whether additional follow-up or withdrawal was warranted. For the follow-up assessment, new instances were enumerated using the same severity definitions, with post-LIFU visit ratings serving as the reference baseline.

#### Tolerability

At least 30 minutes after LIFU or sham sonication, participants completed a three-item experience assessment rated on a 7-point Likert scale (0 = strongly disagree, 1 = disagree, 2 = somewhat disagree, 3 = neutral, 4 = somewhat agree, 5 = agree, 6 = strongly agree), comprising three tolerability items: ‘I felt uncomfortable during the stimulation,’ ‘I felt comfortable during the stimulation,’ and ‘I would recommend participation in this study to others’. Ratings from each LIFU and sham visit were compared using a two-tailed paired Wilcoxon signed-rank test (n=13 paired observations) for each item. Statistical significance was set at α = 0.05

#### Blinding

Participants were blinded to condition assignment throughout the study. The allocation sequence was generated prior to enrollment, stored separately from the research team, and not revealed until all data collection was complete. To maintain blinding to which intervention was delivered, participants were informed that the scheduling of the dedicated safety imaging visit was determined by staff availability. Research staff administering LIFU were aware of condition assignment but were not involved in data analysis. While this minimized observer bias in analysis, it remained possible that administering researchers could have inadvertently conveyed condition information to participants. The three blinding questions were: ‘I could hear the LIFU stimulation,’ ‘I could feel the LIFU stimulation,’ and ‘I believe I experienced LIFU stimulation.’ Ratings for each blinding item were compared between LIFU and sham conditions using a two-tailed paired Wilcoxon signed-rank test (n=13 paired observations). Statistical significance was set at α = 0.05.

#### Auditory Masking

LIFU transducers can produce audible artifacts at the pulse repetition frequency, which falls within the human audible range. To eliminate this potential confound [91,92] continuous acoustic masking was implemented throughout each experimental visit. Participants wore disposable earbuds connected to a tablet running a commercially available white noise application (White Noise Baby Sleep Sounds, AMICOOLSOFT). A multitone noise, previously demonstrated to effectively mask ultrasound-related auditory artifacts [91], was generated and applied across all participants. Volume was set to a comfortable level that eliminated ambient sounds, typically averaging 70–75 dB. Successful masking was confirmed by speaking to participants outside their visual field prior to and during the visit.

### Quantitative Sensory Testing – CPM and TSP

All QST was conducted using a thermal cutaneous stimulator system (TCS, QST.lab, Strasbourg, France) equipped with a contact Peltier thermode (T03, QST.lab, Strasbourg, France). The TCS/T03 system is capable of rapid temperature ramps (ramp-up: 70 °C/s; ramp-down: up to 300 °C/s) and has been validated for the experimental assessment of pain processing in both healthy [93–95] and clinical populations [96].

#### Participant preparation

Prior to formal testing, participants were seated comfortably with the right hand supported and the dorsum facing upwards. The thermode was set to a baseline temperature of 32 °C and placed on the radial aspect of the dorsum of the right hand (corresponding to the C6 dermatome). To familiarize participants with the thermode sensation and minimize startle or novelty effects, temperature was gradually increased at a rate of 1 °C/s until the participant reported a sensation of warmth or heating.

#### Pain thresholding

Participants were individually thresholded to a 6/10 pain intensity using a 0-10 numerical rating scale (NPRS), where 0 = “no pain,” 1-3 = “mild pain,” 4-6 = “moderate pain,” 7-9 = “severe pain,” and 10 = “worst pain imaginable.” Thresholding began 5 °C above the temperature at which warmth was first detected. Three trapezoidal heat stimuli were delivered in series, each consisting of a ramp-up phase (70 °C/s), a brief plateau, and a ramp-down phase (300 °C/s), with a total stimulus duration of 1.8 s. Verbal pain ratings were recorded following each stimulus. Temperature was increased in 1 °C increments until an average pain rating of 6/10 was achieved across the three stimuli. This temperature was used as the individualized stimulus intensity for the TSP protocol.

#### Temporal summation of pain (TSP)

Following thresholding, participants received a train of 10 consecutive heat stimuli delivered at their individualized target temperature. Each stimulus had a total duration of 1.8 s using the same trapezoidal waveform described above, with plateau duration varying slightly across participants due to individual differences in target temperature, averaging approximately 1.4 s. Stimuli were delivered at a fixed inter-stimulus interval (ISI) of 2 s, with the thermode returning to 40 °C between stimuli. Participants verbally rated the perceived pain intensity of each stimulus using the NPRS immediately following delivery. The TSP procedure was performed once prior to LIFU or sham intervention and once following intervention.

TSP was quantified for each visit as the maximum increase in pain rating within the 10-pulse heat stimulation train relative to the first valid rating [22,97]. Specifically, the first valid rating was subtracted from the maximum valid rating observed across the train. If no rating exceeded the first valid rating, a value of zero was assigned to indicate no measurable TSP [22]. A valid rating was defined as one in which the participant tolerated the full heat stimulus and provided a numeric pain rating. Participants were required to have a minimum of 6 valid ratings in both the pre- and post- sonication conditions to be included in the TSP analysis; those who did not meet this threshold due to heat stimulus intolerability or multiple missed ratings were excluded. For each condition, a TSP change score (ΔTSP) was calculated by subtracting the pre-sonication TSP value from the post-sonication TSP value. LIFU and sham ΔTSP change scores were compared using a two-tailed paired Wilcoxon signed-rank test. Statistical significance was set at α = 0.05.

#### Conditioned Pain Modulation (CPM)

CPM was assessed using a heterotopic heat-cold pain paradigm [98]. Baseline heat pain thresholds were first obtained on the dorsum of the right hand using a slowly ramping contact heat stimulus (TCS/T03 system, QST.lab, Strasbourg, France). Temperature increased continuously at a rate of 1 °C/s from a baseline of 32 °C until participants indicated a pain intensity of 6/10 via button press, at which point the temperature immediately returned to baseline. This procedure was repeated three times, and the average temperature at button press was recorded as the baseline heat pain threshold.

The contact heat stimulus on the right hand served as the test stimulus (TS), and the conditioning stimulus (CS) was a left-hand cold-water immersion. Following baseline TS thresholding, participants immersed their left hand up to the wrist in an ice-water bath maintained at 8 °C for 1 minute. Immediately following hand removal, TS was repeated on the dorsum of the right hand using the identical ramping procedure, again averaging across three trials to obtain the post-conditioning threshold. The CPM procedure was performed once prior to LIFU and once following LIFU.

CPM was quantified as the mean post-conditioning TS, computed as the average of three TS trials administered immediately following CS removal [98]. For each condition (LIFU and sham), a CPM change score (ΔCPM) was calculated by subtracting the pre-sonication thresholds from the post-sonication thresholds, such that positive values reflected greater pain inhibition. LIFU and sham ΔCPM change scores were compared using a two-tailed paired Wilcoxon signed-rank test (N = 13). Statistical significance was set at α = 0.05.

## RESULTS

### Patient Characteristics

Thirteen of nineteen participants with a confirmed FM diagnosis completed all study visits (mean age = 43.1 ± 13.2 years; range 24–65; 12 female, 1 male; 1 Latinx, 12 White). N = 6 did not proceed to formal LIFU or sham intervention visits: one was unable to complete the required structural MRI and five were lost to follow-up due to scheduling/availability conflicts. Patient characterization is summarized in **Table 1**. In brief, overall fibromyalgia impact, assessed with the FIQR, fell in the moderate-to-severe range (53.9 ± 16.5; range 30.3–80.5). On the PROMIS framework, in which T-scores are standardized to the U.S. general population (mean = 50, SD = 10), participants showed reduced physical function (T = 35.5 ± 3.0; 1.45 SD below the population mean) and elevated nociceptive pain quality (T = 67.6 ± 8.2; 1.76 SD above the population mean), alongside moderate pain interference (PEG = 5.6 ± 1.9). Pain catastrophizing was in the moderate range (PCS = 25.4 ± 10.0). Group means on the brief screening measures were 2.6 ± 1.9 for anxiety (GAD-2) and 2.2 ± 1.7 for depression (PHQ-2). 5 of 13 participants screened positive for anxiety (GAD-2 ≥ 3) and 5 of 13 screened positive for depression (PHQ-2 ≥ 3), consistent with the established comorbidity of mood symptoms in FM [29,43]. On the TAPS-2, no participant exceeded the threshold for higher-risk substance use. Most participants reported no past-3-month use across substance classes. Nonzero scores were most common for cannabis (8/13; 7 scored 1, 1 scored 2) and alcohol (5/13; 2 scored 1, 3 scored 2), while one participant scored 1 for tobacco. No participants endorsed heroin, opioid misuse, sedative misuse, or stimulant misuse.

**Table 1.** Baseline clinical characterization of the sample (N = 13).

| Measure | Instrument | Mean ± SD (Range) |
| --- | --- | --- |
| Age (years) | — | 43.1 ± 13.2 (24–65) |
| Sex | — | 12 F / 1 M |
| Race/ethnicity | — | 12 White, 1 Latinx |
| Fibromyalgia impact | Fibromyalgia Impact Questionnaire – Revised (FIQR) | 53.9 ± 16.5 (30.3–80.5) |
| Physical function | PROMIS Physical Function Short Form (6-item) | 35.5 ± 3.0 (30.5–41.3) |
| Sleep disturbance | PROMIS Sleep Disturbance Short Form (6a) | 39.9 ± 4.7 (34.2–51.3) |
| Nociceptive pain quality | PROMIS Adult v2.0 Nociceptive Pain Quality (5a) | 67.6 ± 8.2 (56.8–82.9) |
| Neuropathic pain quality | PROMIS Adult v2.0 Neuropathic Pain Quality (5a) | 53.0 ± 11.7 (41.3–82.9) |
| Pain catastrophizing | Pain Catastrophizing Scale (PCS) | 25.4 ± 10.0 (7–44) |
| Pain interference / impact | PEG (Pain, Enjoyment, General Activity) | 5.6 ± 1.9 (3–9) |
| Anxiety symptoms | GAD-2 | 2.6 ± 1.9 (0–6) |
| Depressive symptoms | PHQ-2 | 2.2 ± 1.7 (0–5) |
| Substance use risk | TAPS-2 | See text |
| Current CNS-active medications | Self-report (safety screening) | 10 (77%) |
| Current pregabalin/gabapentinoid | Self-report (safety screening) | 4 (31%) |
| Current opioid analgesic use | Self-report (safety screening) | 1 (8%) |
| Cardiovascular comorbidity* | Self-report (safety screening) | 4 (31%) |
| Other medical comorbidities† | Self-report (safety screening) | See footnote |

Note. Values are mean ± SD (range) for continuous variables, n / total for categorical variables, or n (%) for binary indicators. PROMIS T-scores are standardized to the U.S. general population (mean = 50, SD = 10). CNS = central nervous system. TAPS-2 = Tobacco, Alcohol, Prescription Medication, and Other Substance Use Tool.
\* Cardiovascular comorbidity defined as current use of antihypertensive or cardiac medication (antihypertensives, beta-blockers, anticoagulants).
† Additional comorbidities reported at screening: hypothyroidism (n = 2), endometriosis (n = 1), migraine (n = 1), peripheral neuropathy (n = 1).

Ten of 13 participants (77%) were taking at least one CNS-active medication at the time of enrollment, including gabapentinoids (4 of 13; 31%) and opioid analgesics (1 of 13; 8%). Cardiovascular comorbidity, defined as current use of antihypertensive, beta-blocker, or anticoagulant medication, was present in 4 of 13 participants (31%). Additional comorbidities reported at screening included hypothyroidism (n = 2), endometriosis (n = 1), migraine (n = 1), and peripheral neuropathy (n = 1). See **Table 1**.

### Acoustic Modeling and Target Engagement

Individual-subject acoustic simulations confirmed successful placement of the acoustic focus on or near the left dorsal agranular insula (lh-dAI) target in all 13 participants (**Supplemental Table 1, Supplemental Fig. 2**). Across the cohort, the mean peak intracranial pressure was 76.1 ± 25.6 kPa. Within the target ROI (lh-dAI; mean volume, 1,075 ± 119 mm³), the mean peak pressure was 56.5 ± 26.9 kPa and the mean voxel-wise pressure was 17.9 ± 6.8 kPa. The peak pressure within the ROI reached 73.8 ± 20.1% of the intracranial maximum and exceeded 50% in 11 of 13 participants, indicating that therapeutically relevant acoustic energy reached the target in most cases. The substantial inter-individual variability across these pressure measurements reflects differences in skull morphology that affects ultrasound wave propagation.

### Thermal Modeling

Thermal modeling under conservative assumptions (1 MPa input) predicted a maximum brain tissue temperature rise of 0.4°C (peak absolute temperature 37.4°C) at the acoustic focus, with the highest temperature occurring in cortical bone at 37.8°C (Δ0.8°C above baseline). All simulated temperature increases remained well below the 2°C safety limit recommended by the ITRUSST consensus guidelines [12].

### Neuroradiology Review

Pre- and post-LIFU structural MRI for all 13 participants were reviewed by a board-certified neuroradiologist (D.C.K.); per-subject neuroradiological impressions are summarized in Supplemental Table 2. No participant demonstrated new evidence of edema, microhemorrhage, acute ischemia, or white matter injury on the post-LIFU scan relative to baseline, and no stopping criteria were triggered. All findings noted at baseline were classified as pre-existing and stable on post-LIFU imaging.

Age-typical leukoaraiosis consistent with chronic microvascular ischemic demyelination was identified in 5 of 13 participants and was unchanged on the post-LIFU scan in each case. Benign incidental intracranial findings of no clinical significance, present at baseline and unchanged across visits, were reported in 7 of 13 participants and included a prepontine cystic lesion favored to represent ecchordosis physaliphora (n = 1), a pineal cyst (n = 1), choroid plexus xanthogranuloma (n = 2), an isolated focus of perivascular gliosis (n = 1), and prominent perivascular spaces (n = 2). In one participant, minimal leukoaraiosis was better delineated on the post-LIFU scan than at baseline; the neuroradiologist attributed this difference to technical factors rather than interval change.

### Quantitative Analysis of FLAIR Imaging for Structural Changes

A quantitative voxel-wise analysis of pre- and post-LIFU FLAIR was performed as a complement to the clinical radiological assessment. The relative FLAIR (rFLAIR) framework, in which each voxel’s FLAIR signal is referenced to a contralateral homologous control region, provides an objective per-voxel measurement that is sensitive to subtle signal changes below the threshold of qualitative inspection and enables formal group-level statistical inference. The analytic pipeline applied to a representative single participant is shown in **Fig. 2**; equivalent panels for each cohort participant are provided in **Supplemental Fig. 1**.

**Figure 2.**
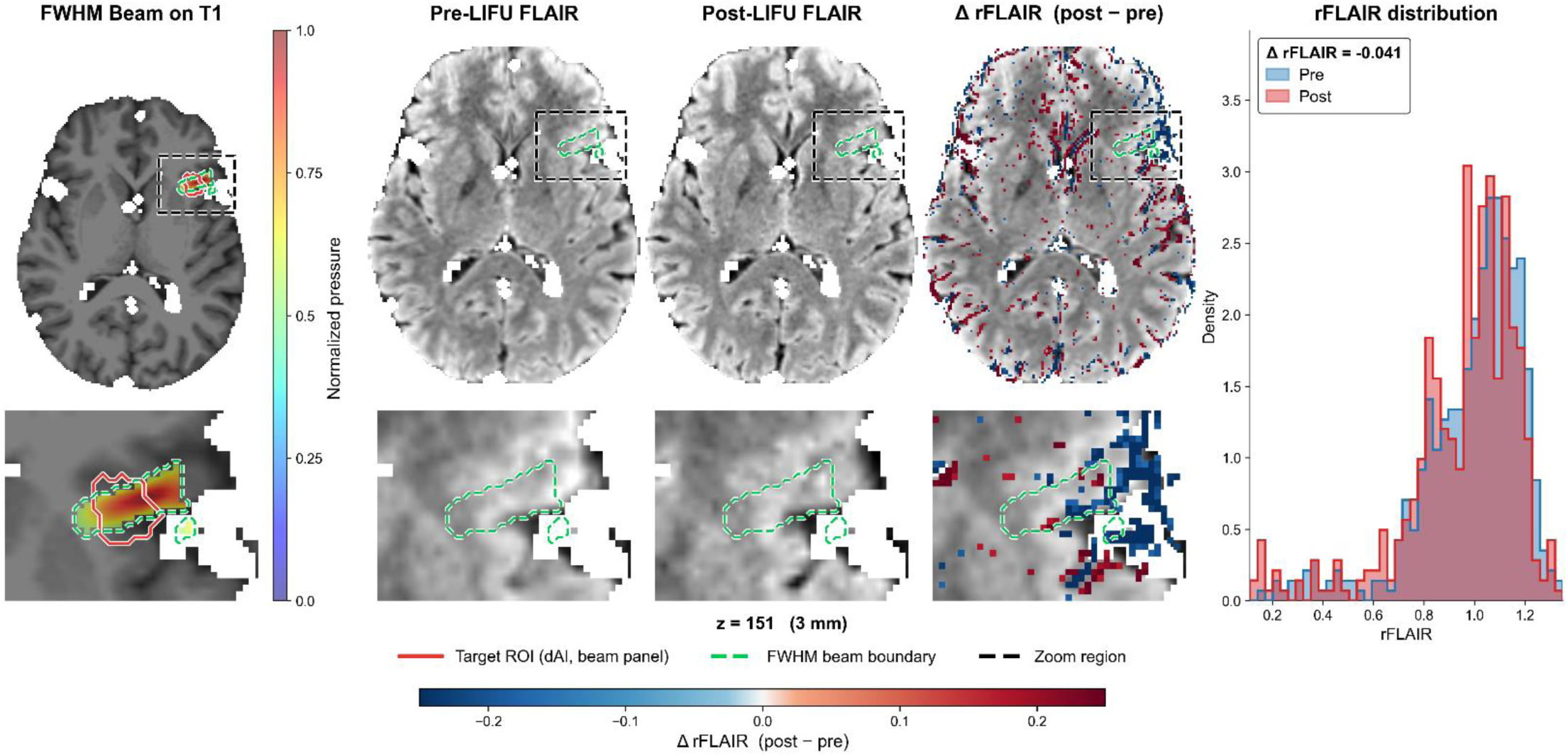
Voxel-wise rFLAIR analysis in a representative single participant. **A.** Normalized intracranial acoustic pressure field (jet colormap) overlaid on T1 MRI, with the left dorsal agranular insula (lh-dAI) target outlined in solid red and the full-width at half-maximum (FWHM) LIFU beam boundary (voxels ≥ 50% of maximum pressure) in dashed green. **B.** From left to right: pre-LIFU FLAIR, post-LIFU FLAIR, voxel-wise ΔrFLAIR overlaid on the pre-LIFU FLAIR image (red, signal increase; blue, decrease post-LIFU), and normalized histograms of within-FWHM beam rFLAIR for the pre- (blue) and post-LIFU (red) sessions with within-subject mean ΔrFLAIR shown in the upper-left. In both **A** and **B**, the upper image shows the full axial slice and the lower image a zoomed inset at the LIFU target (location marked by the dashed rectangle in the upper image); slice z-coordinate is indicated beneath. The horizontal colorbar at the bottom spans the ΔrFLAIR scale in the third column of **B**.

Across the cohort (*n* = 13), the FWHM intracranial LIFU beam volume averaged 634 ± 401 mm³ (range, 192–1,650 mm³) and the contralateral right dorsal agranular insula (rh-dAI) reference ROI averaged 1,075 ± 119mm³ (range, 901–1,261 mm³), at 1 mm³ isotropic resolution in each participant’s T1-native space (**Table 2**). Mean pre-LIFU rFLAIR within the beam volume was 0.93 ± 0.18 (mean ± s.d. across participants; range, 0.35–1.09), and mean post-LIFU rFLAIR was 0.93 ± 0.19 (range, 0.35–1.14), yielding a within-subject change of ΔrFLAIR = +0.002 ± 0.025 (range, −0.041 to +0.046; 95% CI, [−0.011, +0.015]) (**Tables 2** and **3**). This change did not differ significantly from zero (paired two-sided *t*-test: *t*(12) = 0.30, *P* = 0.769; Cohen’s *d*_z = 0.08). A Wilcoxon signed-rank test, used as a check against non-normality of the difference distribution given the small sample size, yielded an equivalent conclusion (*W* = 44.0, *P* = 0.946) (**Table 3**). At the individual level, no participant’s mean post-LIFU beam rFLAIR approached 1.15 (**Table 2**), the conventional threshold used to identify radiologically positive FLAIR lesions in stroke imaging [53,99].

**Table 2.** Per-subject rFLAIR in the FWHM beam volume, pre- and post-LIFU.

| Subject | FWHM<br>Beam (vox) | ROI (vox) | Pre (mean ± SD) | Post (mean ± SD) | ΔrFLAIR |
| --- | --- | --- | --- | --- | --- |
| S01 | 241 | 1117 | 0.97 ± 0.25 | 0.98 ± 0.25 | +0.011 |
| S02 | 429 | 1112 | 1.05 ± 0.16 | 1.07 ± 0.16 | +0.015 |
| S03 | 584 | 1261 | 0.98 ± 0.23 | 0.98 ± 0.22 | -0.004 |
| S04 | 1650 | 922 | 1.09 ± 0.17 | 1.14 ± 0.18 | +0.046 |
| S05 | 515 | 1203 | 0.98 ± 0.18 | 1.02 ± 0.18 | +0.040 |
| S06 | 276 | 901 | 0.35 ± 0.39 | 0.35 ± 0.40 | -0.007 |
| S07 | 476 | 986 | 0.98 ± 0.23 | 0.94 ± 0.25 | -0.041 |
| S08 | 192 | 1159 | 0.93 ± 0.18 | 0.92 ± 0.17 | -0.014 |
| S09 | 469 | 1228 | 1.03 ± 0.20 | 1.01 ± 0.20 | -0.016 |
| S10 | 929 | 1104 | 0.85 ± 0.17 | 0.88 ± 0.19 | +0.026 |
| S11 | 954 | 1028 | 0.97 ± 0.14 | 0.95 ± 0.13 | -0.020 |
| S12 | 967 | 1007 | 0.91 ± 0.16 | 0.90 ± 0.17 | -0.015 |
| S13 | 554 | 941 | 0.93 ± 0.19 | 0.94 ± 0.19 | +0.007 |
| **Mean** | **634** | **1075** | **0.93 ± 0.20** | **0.93 ± 0.21** | **+0.002** |
| **SD** | **401** | **120** | **0.18 ± 0.07** | **0.19 ± 0.07** | **0.025** |

*Note. Pre/Post rFLAIR values are mean ± SD across voxels within each subject's FWHM beam volume. ΔrFLAIR = Post - Pre. Beam (vox) and ROI (vox) report the number of voxels in the FWHM beam volume and the dAI reference ROI, respectively. The final two rows give the across-subject mean and SD of each column (the entries in the Pre/Post columns show the mean and SD of the per-subject means and of the per-subject SDs).*

**Table 3.**
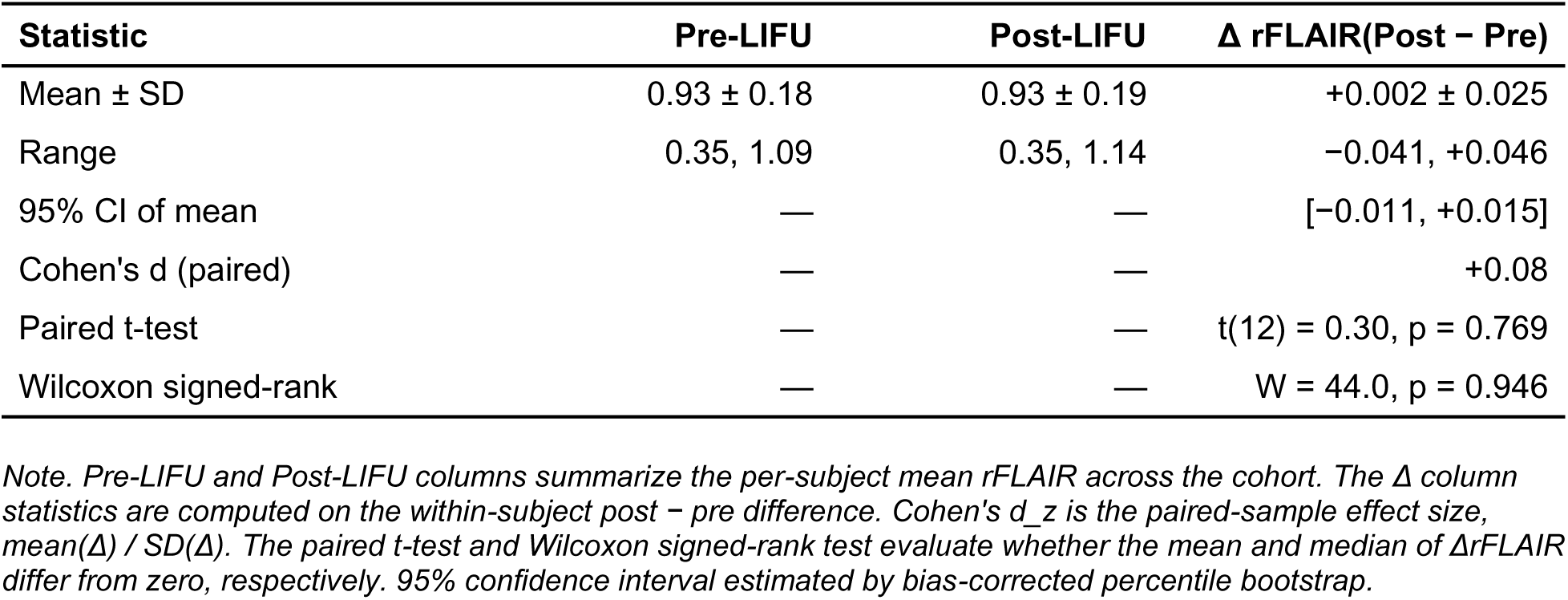
Group-level rFLAIR statistics within the FWHM beam volume (N = 13).

| Statistic | Pre-LIFU | Post-LIFU | $\Delta$ rFLAIR(Post – Pre) |
| --- | --- | --- | --- |
| Mean ± SD | 0.93 ± 0.18 | 0.93 ± 0.19 | +0.002 ± 0.025 |
| Range | 0.35, 1.09 | 0.35, 1.14 | –0.041, +0.046 |
| 95% CI of mean | — | — | [–0.011, +0.015] |
| Cohen's d (paired) | — | — | +0.08 |
| Paired t-test | — | — | t(12) = 0.30, p = 0.769 |
| Wilcoxon signed-rank | — | — | W = 44.0, p = 0.946 |

*Note. Pre-LIFU and Post-LIFU columns summarize the per-subject mean rFLAIR across the cohort. The $\Delta$ column statistics are computed on the within-subject post – pre difference. Cohen's $d_z$ is the paired-sample effect size, $\text{mean}(\Delta) / \text{SD}(\Delta)$ . The paired t-test and Wilcoxon signed-rank test evaluate whether the mean and median of ΔrFLAIR differ from zero, respectively. 95% confidence interval estimated by bias-corrected percentile bootstrap.*

Taken together, the group-level null effect, and the absence of any individual participant exceeding the conventional FLAIR-positivity threshold indicate that single-session LIFU stimulation of the lh-dAI did not induce a detectable acute structural FLAIR signature within the LIFU stimulated tissue.

#### Report of Symptoms

Pre-visit baseline. Pre-visit symptom burden was comparable across both LIFU and sham conditions: 7 of 13 participants reported at least one symptom at moderate or severe intensity prior to both the LIFU and sham visits (**Figure 3**). The most frequently reported pre-visit symptoms were neck pain (LIFU: 9/13, sham: 9/13), sleepiness (LIFU: 8/13, sham: 10/13), forgetfulness (LIFU: 8/13, sham: 7/13), anxiousness (LIFU: 7/13, sham: 6/13), and attention difficulties (LIFU: 5/13, sham: 8/13). Individual-level severity profiles and group-level distributions are shown in **Figure 3**.

**Figure 3.**
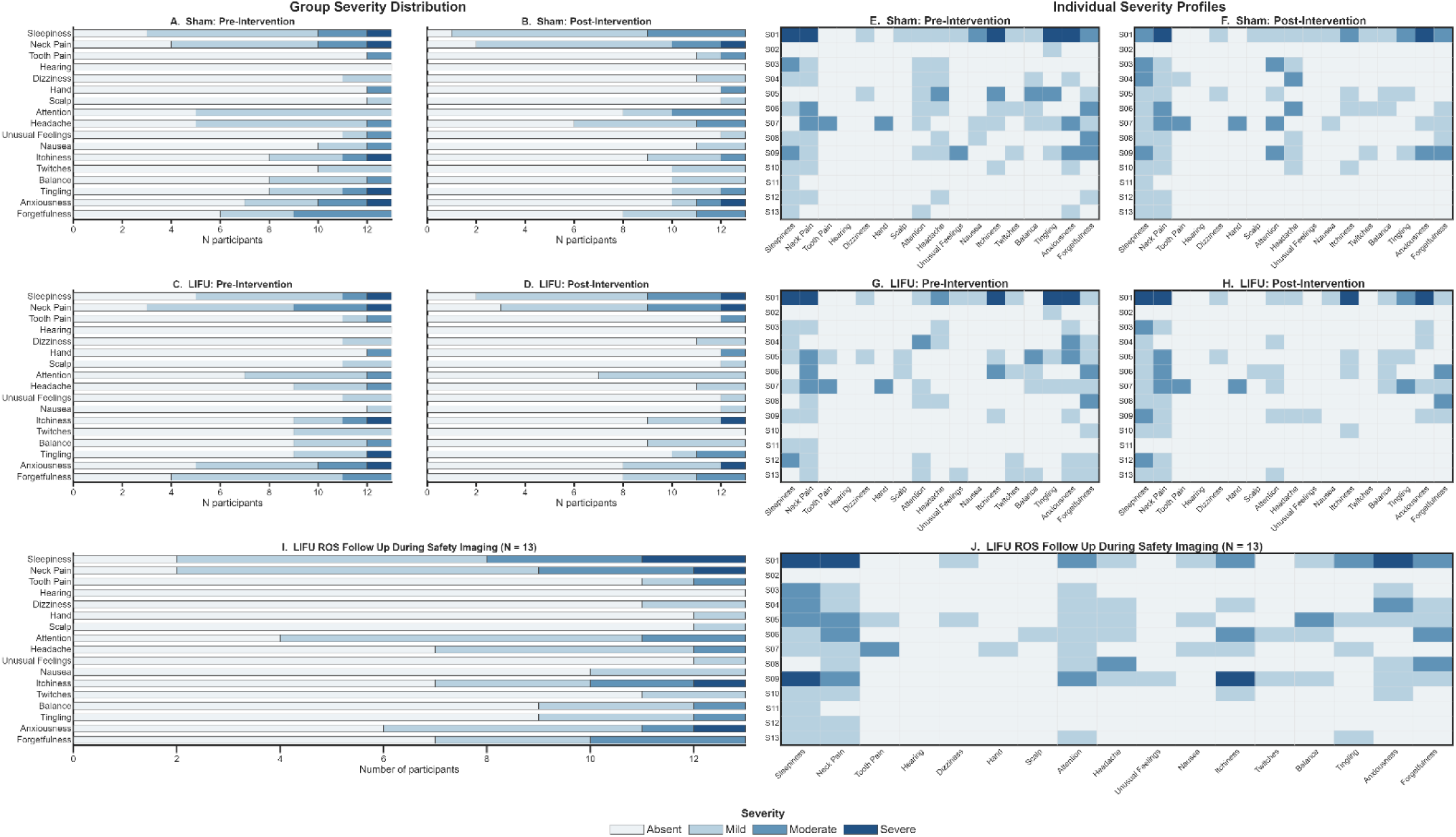
Symptom severity distributions. Symptom severity was assessed on a 4-point ordinal scale (absent, mild, moderate, severe) at three timepoints: before sonication (pre-intervention), at least 30 minutes after sonication (post-intervention), and at 24–72 hours after active LIFU sonication (follow-up). Left panels (A–D, I) show group-level stacked bar distributions, where bar length indicates the number of participants reporting each severity level and stacked segments are color-coded by severity (absent: near-white; mild: light blue; moderate: medium blue; severe: dark navy). Right panels (E–H, J) show individual-level severity heatmaps, where each row corresponds to one participant (S01–S13) and each column to one symptom domain; cell color reflects that participant’s severity rating using the same four-level color palette. **A.** Sham session, pre-intervention. **B.** Sham session, post-intervention. **C.** LIFU session, pre-intervention. **D.** LIFU session, post-intervention. **E.** Sham session, pre-intervention (individual profiles). **F.** Sham session, post-intervention (individual profiles). **G.** LIFU session, pre-intervention (individual profiles). **H.** LIFU session, post-intervention (individual profiles). **I.** LIFU follow-up at safety imaging visit (group distribution). **J.** LIFU follow-up at safety imaging visit (individual profiles).

Within-visit change. No participant experienced a new severe or new moderate symptom elevation following either LIFU or sham. Following LIFU, 6 of 13 participants showed worsening in at least one symptom domain. Sleepiness was worsened in 4 participants (2 absent to mild, 2 mild to moderate), balance in 1 (absent to mild), attention in 1 (absent to mild), and tingling in 1 (mild to moderate). Ten of 13 participants showed improvement in at least one domain following LIFU: headache improved in 4 participants (3 mild to absent, 1 moderate to mild), forgetfulness in 4 (all mild to absent), anxiousness in 4 (2 moderate to mild, 2 mild to absent), and twitches in 3 (all mild to absent). Following sham, 8 of 13 participants showed worsening in at least one domain: headache in 3 (2 mild to moderate, 1 absent to mild), attention in 3 (all mild to moderate), sleepiness in 3 (1 mild to moderate, 2 absent to mild), and neck pain in 2 (both absent to mild). Eleven of 13 participants showed improvement in at least one domain following sham: forgetfulness in 4, anxiousness in 4, and tingling in 4. Within-visit change for all symptom domains across both conditions is shown in **Figure 4 (A & B)**. No stopping criteria were triggered in either LIFU or sham condition, no serious adverse events occurred, and no participants were withdrawn from the study.

**Figure 4.**
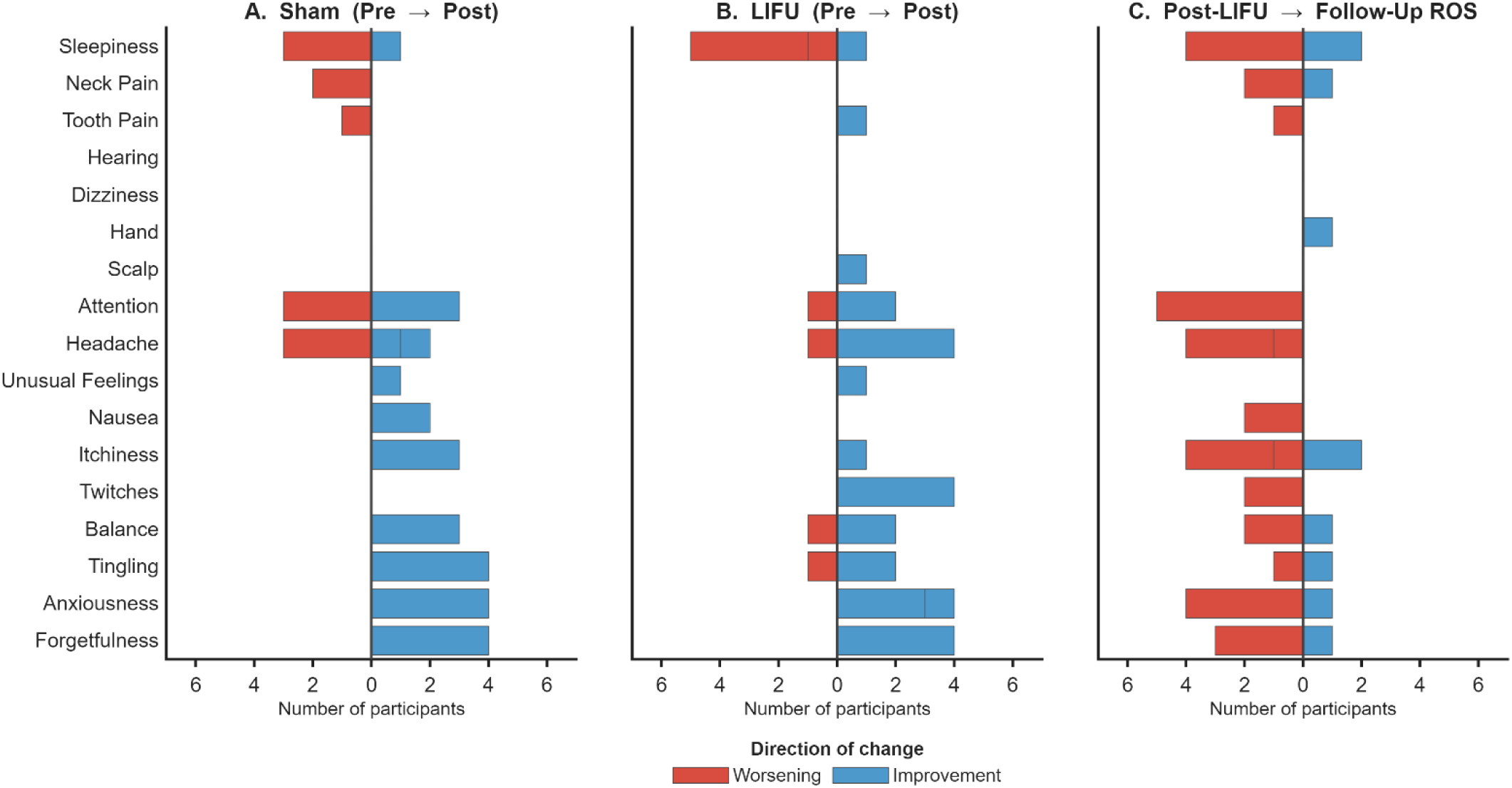
Change in symptom severity following LIFU and sham sonication. Change scores were calculated separately for each of 17 symptom domains rated on a 4-point ordinal scale (absent, mild, moderate, severe). For Panels A and B, change = pre-intervention rating minus post-intervention rating such that blue bars indicate improvement and red bars indicate worsening. For Panel C, change = post-LIFU rating minus follow up visit (24 – 72 hours) rating, such that blue bars indicate symptom improvement, and red bars indicate symptom worsening or new onset over that time interval. Bar length indicates the number of participants. **A.** Sham within-session change. **B.** LIFU within-session change. **C.** 24 to 72 hours follow-up change from post-LIFU. All panels are N = 13.

Follow-up assessment. A follow-up ROS was administered at the dedicated safety imaging visit (24 – 72 hours) for all 13 participants. At the individual level, 11 of 13 participants showed worsening in at least one symptom domain at follow-up relative to post-LIFU ratings (**Figure 4**). The most common domains that worsened were attention (5/13), headache (4/13), itchiness (4/13), sleepiness (4/13), and anxiousness (4/13). One new moderate instance was observed: headache was absent at the post-LIFU assessment and rated moderate at the follow-up visit in 1 participant. One new severe instance was observed: itchiness was absent at the post-LIFU assessment and rated severe at the follow-up visit in 1 participant. Review of severe itchiness confirmed no clinical intervention was warranted and no participant was withdrawn. Five of 13 participants showed improvement in at least one symptom domain at follow-up (**Figure 4**).

#### Tolerability

Three tolerability items (uncomfortable, comfortable, would recommend) were rated separately following the sham and LIFU visits on a 7-point Likert scale (**Figure 5**).

**Figure 5.**
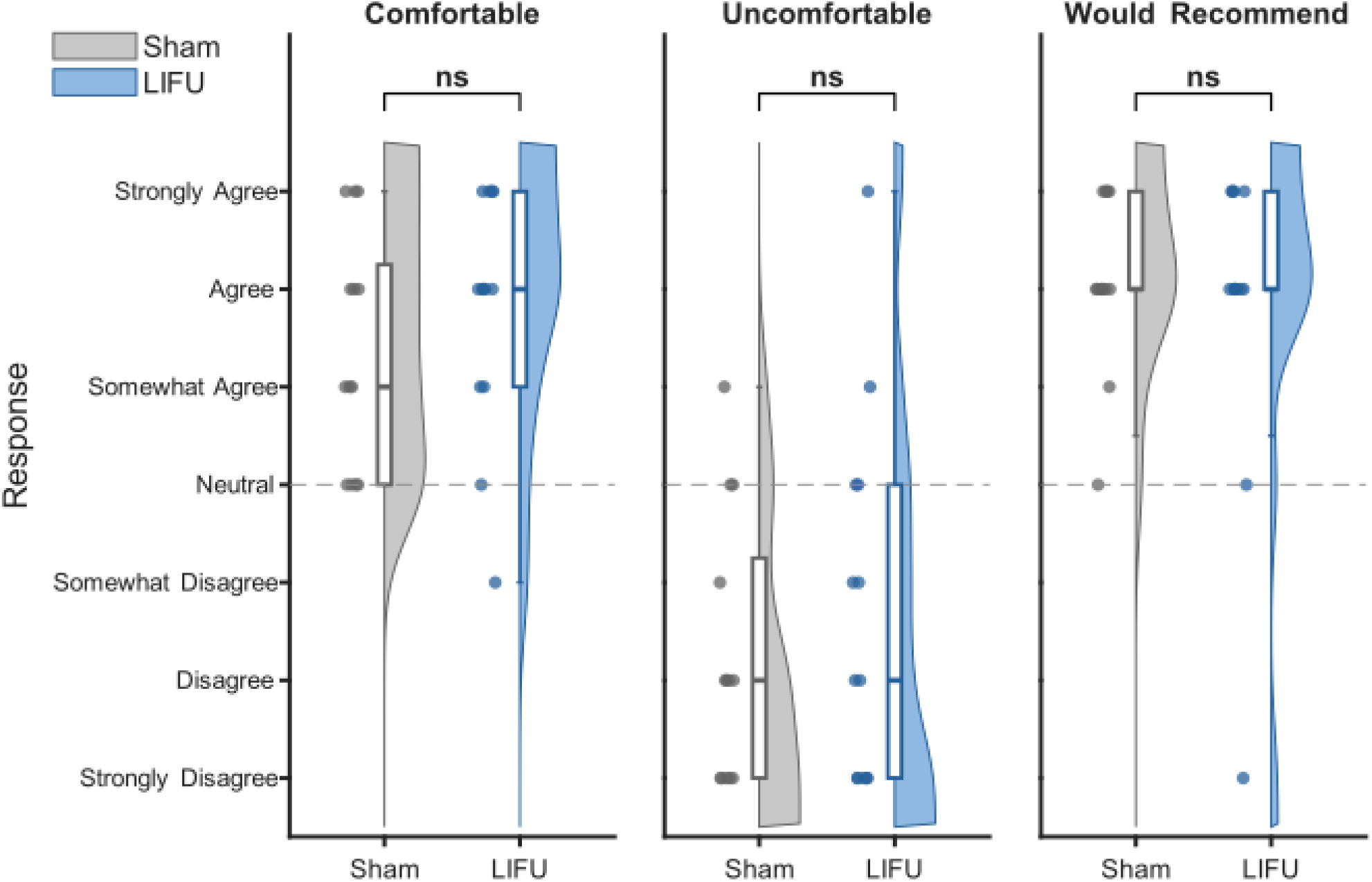
Tolerability ratings. At least 30 minutes after each sonication session, participants rated three tolerability items (’I felt uncomfortable during the stimulation,’ ‘I felt comfortable during the stimulation,’ and ‘I would recommend participation in this study’) on a 7-point Likert scale (0 = strongly disagree, 1 = disagree, 2 = somewhat disagree, 3 = neutral, 4 = somewhat agree, 5 = agree, 6 = strongly agree; full anchors shown on the y-axis). For each item, sham (grey) and LIFU (blue) responses are shown as raincloud plots: the half-violin shows the distribution of ratings (kernel density estimate), the box plot indicates median and interquartile range, and individual participant ratings are overlaid as jittered points. The dashed horizontal line marks the scale neutral midpoint (3). Brackets above each panel show the paired Wilcoxon signed-rank comparison of sham versus LIFU; ns = not significant (p > 0.05).

For ‘I felt uncomfortable during the stimulation,’ mean ratings were 1.23 ± 1.36 (mean ± SD; range 0–4) at the sham visit and 1.69 ± 1.89 (mean ± SD; range 0–6) at the LIFU visit, corresponding in both conditions to a position between ‘disagree’ and ‘somewhat disagree.’ At the sham visit, 10 of 13 ratings were at or below ‘somewhat disagree’ (scores ≤ 2), with the remaining 3 distributed as 2 ratings of ‘neutral’ and 1 of ‘somewhat agree.’ At the LIFU visit, 9 of 13 ratings were at or below ‘somewhat disagree,’ with the remaining 4 distributed as 2 ratings of ‘neutral,’ 1 of ‘somewhat agree,’ and 1 of ‘strongly agree.’ The paired sham-versus-LIFU comparison was not significant (W = 18.5, p = 0.38) (**Figure 5**).

For ‘I felt comfortable during the stimulation,’ mean ratings were 4.38 ± 1.19 (mean ± SD; range 3–6) at the sham visit and 4.77 ± 1.24 (mean ± SD; range 2–6) at the LIFU visit, corresponding in both conditions to a position between ‘somewhat agree’ and ‘agree.’ At the sham visit, 9 of 13 ratings were at ‘somewhat agree’ or higher (scores ≥ 4), with the remaining 4 all rated ‘neutral.’ At the LIFU visit, 11 of 13 ratings were at ‘somewhat agree’ or higher, with the remaining 2 distributed as 1 rating of ‘neutral’ and 1 of ‘somewhat disagree.’ The paired sham-versus-LIFU comparison was not significant (W = 14.5, p = 0.42) (**Figure 5**).

For ‘I would recommend participation in this study,’ mean ratings were 5.08 ± 0.86 (mean ± SD; range 3–6) at the sham visit and 4.77 ± 1.64 (mean ± SD; range 0–6) at the LIFU visit, corresponding in both conditions to a position near ‘agree.’ At the sham visit, 12 of 13 ratings were at ‘somewhat agree’ or higher (scores ≥ 4), with the remaining 1 rated ‘neutral.’ At the LIFU visit, 11 of 13 ratings were at ‘somewhat agree’ or higher, with the remaining 2 distributed as 1 rating of ‘neutral’ and 1 of ‘strongly disagree.’ The paired sham-versus-LIFU comparison was not significant (W = 3.0, p = 1.00) (**Figure 5**).

#### Blinding

Three blinding items (could hear, could feel, experienced LIFU) were rated separately following the sham and LIFU visits on a 7-point Likert scale (Figure 6).

**Figure 6.**
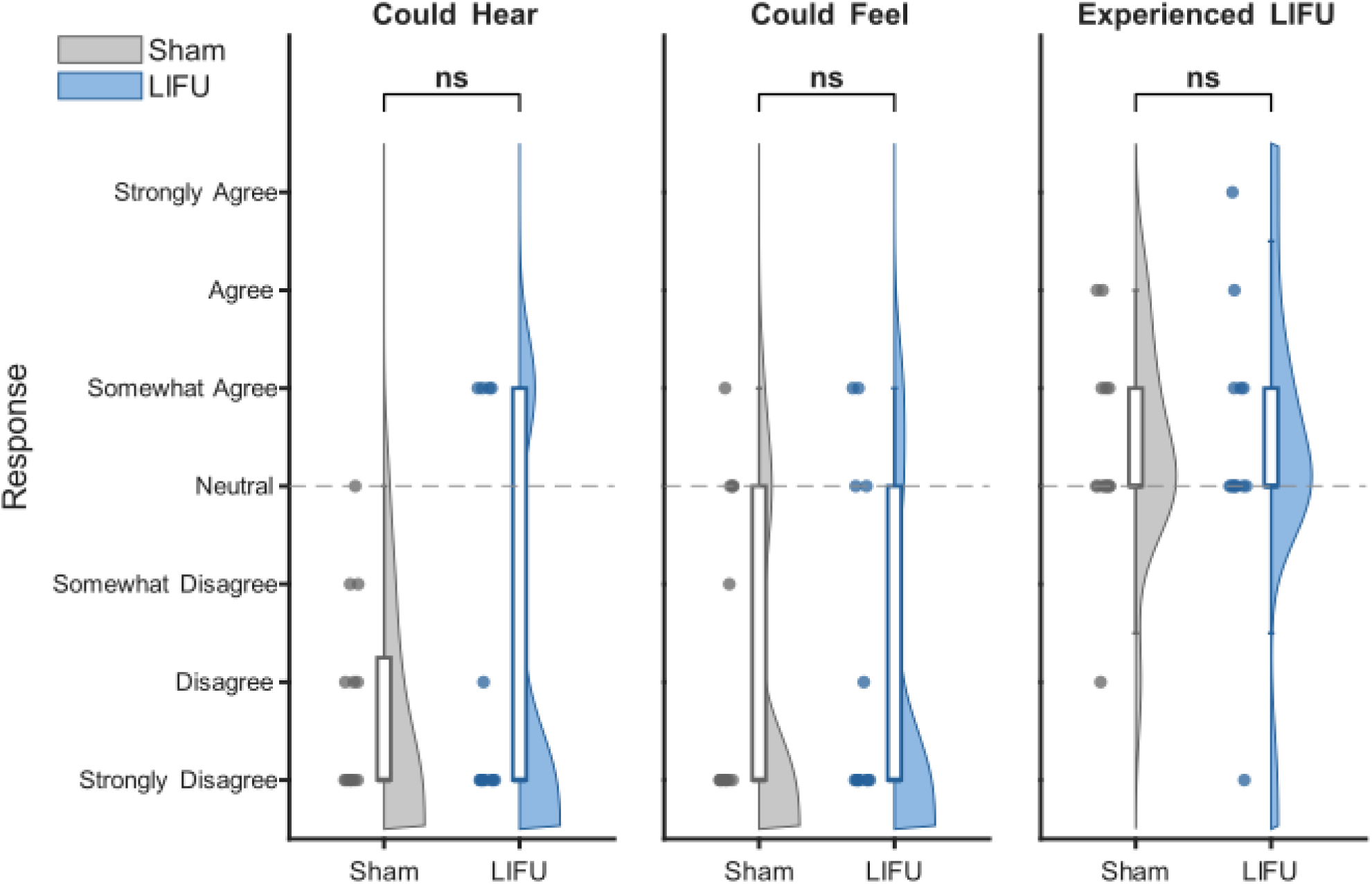
Blinding Ratings. At least 30 minutes after each sonication session, participants rated three blinding items (’I could hear the LIFU stimulation,’ ‘I could feel the LIFU stimulation,’ and ‘I believe I experienced LIFU stimulation’) on a 7-point Likert scale (0 = strongly disagree, 1 = disagree, 2 = somewhat disagree, 3 = neutral, 4 = somewhat agree, 5 = agree, 6 = strongly agree; full anchors shown on the y-axis). For each item, sham (grey) and LIFU (blue) responses are shown as raincloud plots: the half-violin shows the distribution of ratings (kernel density estimate), the box plot indicates median and interquartile range, and individual participant ratings are overlaid as jittered points. The dashed horizontal line marks the scale neutral midpoint (3). Brackets above each panel show the paired Wilcoxon signed-rank comparison of sham versus LIFU; ns = not significant (p > 0.05).

For ‘I could hear the LIFU stimulation,’ mean ratings were 0.77 ± 1.01 (mean ± SD; range 0–3) at the sham visit and 1.31 ± 1.89 (mean ± SD; range 0–4) at the LIFU visit, corresponding in both conditions to a position between ‘strongly disagree’ and ‘disagree.’ At the sham visit, 12 of 13 ratings were at or below ‘somewhat disagree’ (scores ≤ 2), with the remaining 1 rated ‘neutral.’ At the LIFU visit, 9 of 13 ratings were at or below ‘somewhat disagree,’ with the remaining 4 all rated ‘somewhat agree.’ The paired sham-versus-LIFU comparison was not significant (W = 5.5, p = 0.34) (**Figure 6**).

For ‘I could feel the LIFU stimulation,’ mean ratings were 1.15 ± 1.57 (mean ± SD; range 0–4) at the sham visit and 1.15 ± 1.68 (mean ± SD; range 0–4) at the LIFU visit, corresponding in both conditions to a position near ‘disagree.’ At the sham visit, 9 of 13 ratings were at or below ‘somewhat disagree,’ with the remaining 4 distributed as 3 ratings of ‘neutral’ and 1 of ‘somewhat agree.’ At the LIFU visit, 9 of 13 ratings were at or below ‘somewhat disagree,’ with the remaining 4 distributed as 2 ratings of ‘neutral’ and 2 of ‘somewhat agree.’ The paired sham-versus-LIFU comparison was not significant (W = 10.0, p = 1.00) (**Figure 6**).

For ‘I believe I experienced LIFU stimulation,’ mean ratings were 3.38 ± 1.04 (mean ± SD; range 1–5) at the sham visit and 3.38 ± 1.39 (mean ± SD; range 0–6) at the LIFU visit, corresponding in both conditions to a position at ‘neutral.’ Ratings were centered on the scale midpoint in both conditions: at the sham visit, 7 of 13 ratings were at ‘neutral,’ 5 of 13 were above neutral (3 ‘somewhat agree,’ 2 ‘agree’), and 1 of 13 was below neutral (’disagree’); at the LIFU visit, 7 of 13 ratings were at ‘neutral,’ 5 of 13 were above neutral (3 ‘somewhat agree,’ 1 ‘agree,’ 1 ‘strongly agree’), and 1 of 13 was below neutral (’strongly disagree’). The paired sham-versus-LIFU comparison was not significant (W = 22.5, p = 1.00) **(Figure 6**).

### Quantitative Sensory Testing

#### TSP

Two of 13 participants did not provide a sufficient number of valid ratings (≥6 of 10) in at least one of the LIFU or sham intervention visits and were excluded from the TSP analysis. In the remaining 11 participants, mean ± SEM TSP magnitude was 2.73 ± 0.62 before LIFU and 1.45 ± 0.41 after LIFU. In the sham condition, mean ± SEM TSP magnitude was 2.82 ± 0.62 before sham and 1.73 ± 0.47 after sham. The resulting mean ± SEM ΔTSP was −1.27 ± 0.30 following LIFU and −1.09 ± 0.53 following sham (**Figure 7A**). A paired Wilcoxon signed-rank test revealed no significant difference between conditions (W = 11.5, p = 0.797).

**Figure 7.**
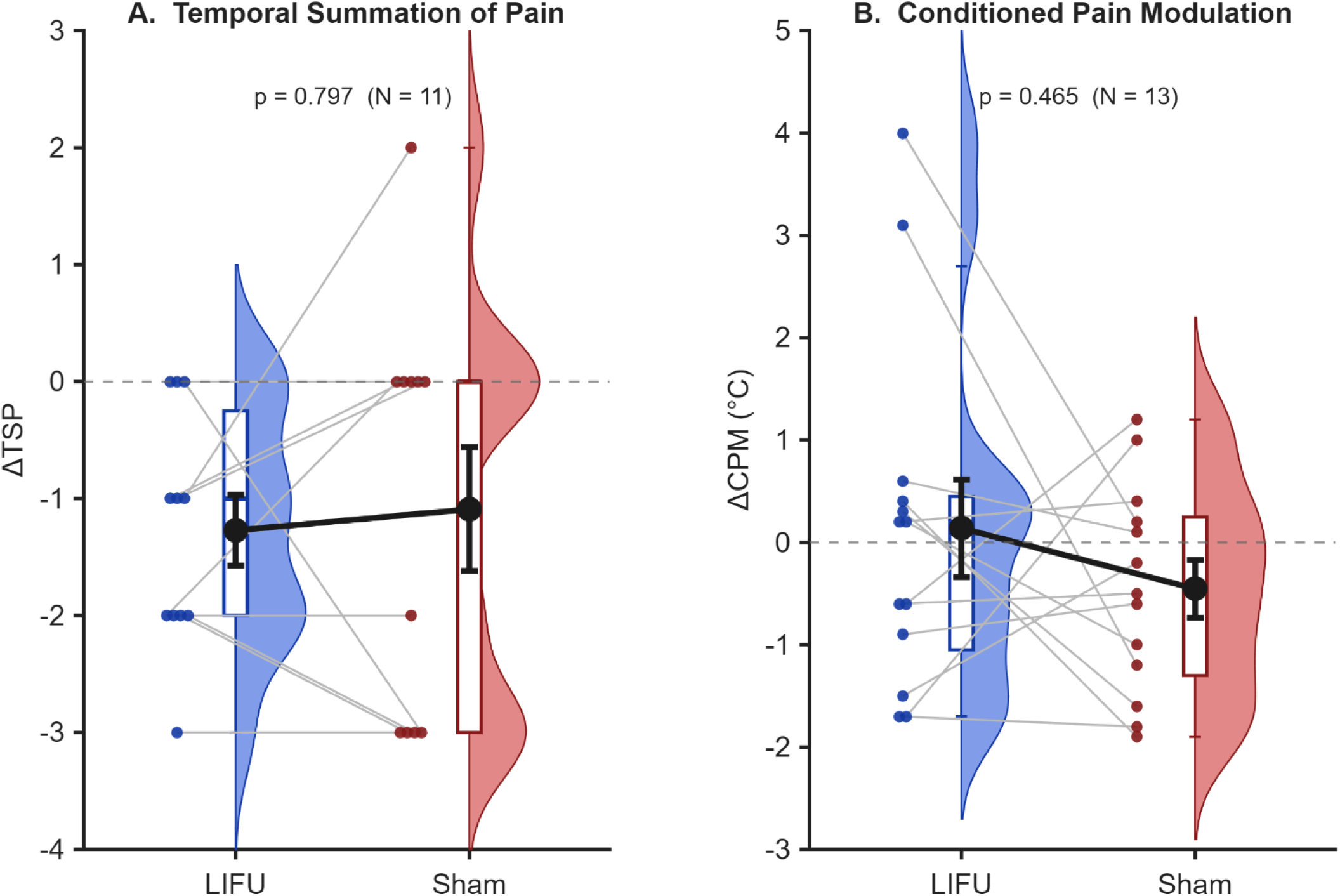
Quantitative Sensory Testing. **A.** Change in temporal summation of pain (TSP) for LIFU (blue) and sham (red) sessions. Group (N = 11) mean ± SEM ΔTSP was −1.27 ± 0.30 following LIFU and −1.09 ± 0.53 following sham. **B.** Change in conditioned pain modulation (CPM) for LIFU (blue) and sham (red). Group (N = 13) mean ± SEM ΔCPM was 0.14 ± 0.48 following LIFU and −0.47 ± 0.28 following sham. In both panels, raincloud plots are shown for LIFU and sham conditions: the half-violin shows the distribution of participant change scores (kernel density estimate), the box plot indicates the median and interquartile range, and filled circles show individual participant change scores. Gray lines connect paired within-participant LIFU and sham values, and black circles with error bars show the group mean ± SEM. The dashed horizontal line indicates no pre-to-post change. Negative ΔTSP values indicate reduced temporal summation following sonication, whereas positive ΔCPM values indicate increased pain threshold following conditioning, consistent with enhanced conditioned pain modulation. LIFU and sham change scores did not differ significantly for either TSP (Wilcoxon signed-rank p = 0.797) or CPM (Wilcoxon signed-rank p = 0.465).

#### CPM

In all 13 participants, the group mean ± SEM ΔCPM (post − pre threshold, °C) was +0.14 ± 0.48 °C following LIFU and −0.45 ± 0.28 °C following sham (**Figure 7B**). A paired Wilcoxon signed-rank test revealed no significant difference between conditions (W = 56.5, p = 0.47). Two participants showed unusually large positive ΔCPM at the LIFU visit (+4.0 °C and +3.1 °C); excluding these participants, the LIFU group mean was −0.48 °C, indicating that the apparent LIFU-side asymmetry was driven by these two responders.

## DISCUSSION

This study evaluated the safety and tolerability of a single 500 kHz, 10-minute application of LIFU targeting the left dorsal anterior insula (dAI) in individuals with fibromyalgia (FM). Across 13 participants who completed the within-subjects sham-controlled crossover, neuroradiological review of post-LIFU structural MRI revealed no new findings of edema, microhemorrhage, acute ischemia, or white matter injury attributable to the intervention. The quantitative voxel-wise FLAIR analysis recapitulated this, demonstrating no evidence of LIFU-induced change at the stimulation site. No serious adverse events were reported and the report of symptoms assessment found similar distributions of symptoms and severity across the LIFU and sham conditions. In addition, participants generally rated the procedure as comfortable and indicated they would recommend participation. Despite this encouraging safety and tolerability profile, LIFU as delivered here, did not result in statistically significant differences in TSP or CPM between LIFU and sham conditions.

### Clinical and Structural Analysis

Qualitative neuroradiological review by a board-certified neuroradiologist identified no evidence of edema, microhemorrhage, acute ischemia, or white matter injury attributable to LIFU on post-LIFU structural imaging. Incidental findings observed in a subset of participants, including age-typical leukoaraiosis, benign cystic lesions, prominent perivascular spaces, and choroid plexus xanthogranulomata, were present at baseline and unchanged on the post-LIFU structural imaging. Quantitative voxel-wise analysis of pre- and post-LIFU FLAIR provided a complementary characterization of signal change within the acoustic focus. The within-subject change in rFLAIR at the stimulation site was small (0.002 ± 0.025). The distribution of ΔrFLAIR values is very close to zero and showed no systematic directional change, with both positive and negative excursions, consistent with physiological measurement noise rather than a directional LIFU effect.

Fluid-attenuated inversion recovery (FLAIR) MRI provides a sensitive marker of structural brain injury by suppressing cerebrospinal fluid signal while preserving sensitivity to T2-prolonging parenchymal abnormalities. Increased FLAIR signal primarily reflects elevations in tissue water content, including vasogenic edema associated with blood–brain barrier disruption and cytotoxic edema reflecting intracellular swelling, both of which prolong T2 relaxation and are well-characterized features of traumatic and non-traumatic brain injury [73,100]. In traumatic brain injury (TBI), FLAIR hyperintensity corresponds to regions of edema, gliosis, and inflammatory response and can detect relatively subtle abnormalities even in mild or diffuse injury states [101–103]. Notably, FLAIR has also demonstrated sensitivity to ultrasound-related tissue effects in clinical contexts, including a reported serious adverse event in a transcranial focused ultrasound study where transient FLAIR hyperintensity was observed within the targeted region [104]. In the present study, the absence of both neuroradiological abnormalities and quantitative FLAIR signal changes indicates no detectable increase in tissue water content within the sonicated volume, suggesting that the LIFU exposure as performed here (10 minutes; 25 – 150 kPa) did not induce edema, blood–brain barrier disruption, or tissue injury at the resolution of clinical MRI.

It is important to place the exposure used here (and the null FLAIR findings) in the context of prior reports of ultrasound neuromodulation and associated imaging and histological findings. In a recent clinical case report, transcranial focused ultrasound delivered at substantially higher acoustic exposures (∼1.3–3.5 MPa) was associated with evolving FLAIR hyperintensities, edema, and microhemorrhages, with imaging abnormalities progressing over days to weeks and ultimately resulting in structural tissue changes [104]. In contrast, human LIFU neuromodulation studies typically operate at substantially lower pressures, often spanning approximately ∼100–700 kPa in situ [16–19,105]. Notably, direct neuroradiological assessment using FLAIR has not been incorporated in these studies, limiting evaluation of subtle tissue responses such as edema. In parallel, converging histological evidence from both human and large-animal studies supports the absence of tissue injury within the low-intensity neuromodulation regime [106,107]. In patients undergoing temporal lobe resection following LIFU exposure, histological analysis revealed no evidence of necrosis, vascular damage, apoptosis, or hemorrhage in sonicated tissue, at exposures corresponding to a reported mechanical index of approximately 2.1 (at 650 kHz), which corresponds to an estimated peak rarefactional pressure on the order of ∼1.5–2.0 MPa [107]. In addition, controlled studies in macaques and sheep have demonstrated no pre-mortem histological abnormalities including no edema, hemorrhage, or apoptotic changes across a broad range of exposures, including in situ peak pressures of approximately ∼0.25–0.9 MPa for neuromodulation and up to ∼1.7–3.6 MPa for brief MR-ARFI applications [106]. In contrast, the ultrasound-associated adverse event described above has been estimated to involve in situ peak negative pressures of approximately ∼1.3–3.5 MPa and elevated mechanical index values exceeding established non-significant risk thresholds, placing it outside the low-intensity neuromodulation regime [108]. In this context, the estimated in-head rarefactional pressures used in the present study (50–125 kPa at 500 kHz) fall at the lower end of the LIFU exposure spectrum and correspond to MI values of approximately 0.07–0.18, well below the MI/MItc ≤ 1.9 threshold considered non-significant risk for cavitation-related mechanical bioeffects [12]. The absence of both neuroradiological abnormalities and quantitative FLAIR signal changes in the present study therefore provides a sensitive, imaging-based confirmation of tissue safety, indicating no detectable edema or structural disruption within the sonicated volume and extending prior histological findings to a clinically accessible MRI-based endpoint.

Thermal simulation performed under conservative assumptions including 65% skull absorption with all absorbed energy converted to heat, predicted a peak brain tissue temperature rise of 0.4°C at the focus and a peak cortical bone rise of 0.8°C, both below the 2°C ITRUSST-recommended limit and below the 39°C absolute-temperature criterion for non-significant thermal risk [12]. However, if heating were sufficient to induce edema, blood–brain barrier disruption, inflammation, or tissue injury, these processes would be expected to increase tissue water content and could manifest as increased FLAIR signal. Thus, the absence of neuroradiological abnormalities and quantitative FLAIR signal increases provides convergent evidence that the present exposure did not produce MRI-detectable thermal or structural tissue effects.

These findings also inform the practical question of whether delayed structural MRI should be required in future LIFU neuromodulation studies. The results here, together with prior human and large-animal histological studies showing no necrosis, apoptosis, hemorrhage, edema, or vascular injury across substantially higher but still controlled neuromodulatory exposures [106,107], suggest that routine delayed FLAIR imaging may have limited yield for low-intensity protocols operating well within established mechanical and thermal safety margins [12,108]. However, the recent report of ultrasound-associated FLAIR hyperintensity following substantially higher exposures underscores that FLAIR is a sensitive and clinically meaningful tool when exposure conditions approach or exceed low-intensity safety regimes.

Although we observed no evidence of FLAIR signal abnormality after low-pressure LIFU exposure in this FM cohort, safety findings from healthy or relatively low-risk samples may not generalize fully to all clinical populations. Neurological disease can alter baseline excitability, vascular integrity, blood–brain barrier function, edema burden, tissue composition, and medication-related seizure threshold, all of which may influence the biological response to non-invasive brain stimulation. This issue is well recognized in TMS safety guidance, where neurological injury, stroke, epilepsy, medications, and other factors are treated as risk modifiers rather than absolute contraindications [109]. For transcranial ultrasound, current biophysical safety consensus similarly frames “nonsignificant risk” thresholds as applying to individuals without contraindications such as compromised thermoregulation or vascular vulnerabilities, or to ultrasound contrast agents; and that mechanical and thermal risk are considered low when MI/MItc and thermal-dose limits are respected [12]. Thus, while our data support the tolerability of low-pressure LIFU in FM under the present parameters, future studies in populations with overt structural lesions, prior stroke, hemorrhage, edema, tumors, demyelination, or vascular abnormalities may warrant additional screening and imaging surveillance. For low-pressure protocols such as the present study, future work may support replacing routine delayed FLAIR with a risk-stratified approach incorporating prospective symptom monitoring, neurological assessment, conservative acoustic modeling, and MRI follow-up only when clinically indicated.

### Report of Symptoms

The primary within-visit safety finding was the absence of new severe symptoms in either the LIFU or sham condition. Per protocol, a new severe symptom was defined as a symptom rated absent at pre-visit baseline reaching severe post-sonication; no such events occurred, and no stopping criteria were triggered in either condition. No new moderate instances were observed in either condition. Symptom changes in both conditions were confined to single-step ordinal shifts in specific domains. A new severe instance of itchiness triggered review, which confirmed no clinical intervention was warranted and no participant was withdrawn. In a single-visit crossover design with a follow-up visit occurring within 24-72 hours post-LIFU, these events are difficult to causally attribute to the intervention, and the worsening across 11 of 13 participants at follow-up is consistent with the day-to-day symptom variability characteristic of FM [110,111].

In general, participants had substantial pre-visit symptom burden that included instances of neck pain, sleepiness, forgetfulness and anxiousness. This is somewhat elevated compared to healthy volunteers [13] however, the change in scores (increase or decrease) across both LIFU and sham conditions demonstrated similar variability to healthy cohorts suggesting a similar report of symptoms safety profile. The present findings are consistent with that profile: no causally attributable severe symptom elevations, despite a markedly greater pre-visit symptom burden and an older, FM patient sample.

In the randomized double-blind sham-controlled crossover trial of Riis et al. (2024) [19], 20 patients with chronic pain, including 10 with FM, received 40 minutes of active or sham FUS targeting the anterior cingulate cortex (ACC) using a 650 kHz phased array delivering an estimated 1 MPa at target. Using the General Assessment of Side Effects (GASE), side effects included headache, fatigue, and musculoskeletal discomfort, that were all rated as mild and resolved within 24 hours. No adverse events were reported and no significant differences between active and sham were observed on any item. Safety assessment in that trial was limited to the GASE recorded at baseline and 24 hours after each treatment session; no post-LIFU structural MRI was acquired, with imaging used only for pre-treatment device registration and target engagement validation. Several design features distinguish that trial from the present study: the target was the ACC rather than the dAI, the sonication duration was four times longer, and the at-target pressure was approximately an order of magnitude higher (∼1 MPa versus 50–125 kPa in-head in the present study) with a corresponding MI of 1.2 versus 0.07–0.18. That both studies found no adverse events and no between-condition differences in symptom burden, despite these implementation differences, indicates that tolerability of single-visit LIFU in chronic pain populations that include patients with FM. The use of different safety instruments across studies, the ROS in the present study and the GASE in Riis et al., limits direct symptom-profile comparison, reinforcing the case for a standardized symptom-monitoring framework across LIFU neuromodulation research as recently proposed [54].

### Tolerability

Participants rated the procedure as ‘comfortable’ and indicated they would recommend participation. Importantly, no differences were observed between LIFU and sham on any tolerability item, no participant prematurely aborted study collection and no participant was withdrawn for tolerability issues. Participants tolerated 10 minutes of continuous transducer-to-scalp contact and sonication without premature termination, establishing a lower-bound duration for single delivery dAI-directed LIFU in FM. How tolerability scales with longer sonication duration remains an open question: sham-controlled trials in chronic pain have employed sonication durations of up to 40 minutes without adverse events or tolerability-related withdrawal [19], but whether therapeutic benefit in FM would require durations beyond those studied here has not been established. These ratings therefore provide a benchmark against which multi-application and longer-duration protocols can be evaluated as the field moves toward clinical application.

Despite the tolerability of LIFU, tolerability of the QST protocol was limited. Two of 13 participants were unable to complete the TSP heat-stimulus sequence due to intolerance of the noxious thermal input and were excluded from analysis, reducing the effective sample size for heat-based outcomes. This finding is consistent with the heightened sensory sensitivity observed in FM [43,44] and highlights a practical limitation of experimental pain paradigms in this population. Future studies should anticipate such attrition and consider incorporating validated clinical pain measures that do not require noxious stimulation.

### Blinding

Blinding integrity is a necessary precondition for causal inference in a sham-controlled design. Ratings for auditory and somatic detection of LIFU were both below neutral and belief ratings clustered near neutral and no LIFU-sham differences were observed on any blinding item. These results indicate that participants could not reliably distinguish active from sham sonication as performed here (and similar to our previous studies in healthy volunteers [20–22]) and highlights the need and use of adequate auditory masking to help prevent revealing allocation [91,92]. The present study however employed a single-blind design in which the administering researcher was aware of condition assignment. Recent work from our group has validated a gel-based acoustic coupling approach that achieves effective double-blinding for LIFU studies [78], and this method should be adopted in future clinical trials where double blinding is desired.

### Relationship of Targeting/Dosing on Quantitative Sensory Testing

TSP and CPM showed no significant differences between LIFU and sham. Interpretation of this null result benefits from a closer consideration of three factors: the functional role of the dAI in pain processing relative to the outcomes measured, the intracranial LIFU exposure, and the geometric overlap of the acoustic focus and the intended target.

The dAI is a mechanistically plausible target in fibromyalgia because FM is not simply a disorder of peripheral nociceptive input, but a condition characterized by central amplification, multisensory hypersensitivity, altered endogenous pain modulation, and abnormal salience/interoceptive processing [23]. Abnormalities in the insula are implicated in the pathophysiology of FM, with evidence for altered activity, structure, connectivity, neurochemistry, and network hub status [28,35–38,44,112]. In particular, the anterior insula appears to acquire abnormal network importance in FM, functioning as a hub/rich-club node in patients but not controls, suggesting that it may disproportionately influence whole-brain information flow relevant to pain, salience, affect, and bodily state monitoring [112]. This makes dAI an attractive target for neuromodulation aimed at reducing the broader symptom complex of FM, including pain amplification, sensory sensitivity, affective distress, fatigue, and interoceptive hypervigilance, rather than only evoked pain sensitivity.

#### Why dAI may not strongly affect TSP

TSP is often treated as a proxy for bottom-up pain facilitation or spinal/ascending central sensitization [55,97]. It is closely linked to C-fiber temporal summation and dorsal horn wind-up, with supraspinal correlates including thalamus, S1/S2, posterior and anterior insula, ACC, SMA, and PFC [42,44]. However, the strongest mechanistic interpretation of TSP remains ascending nociceptive facilitation, and the posterior insula appears more directly related to sensory-discriminative nociceptive coding, including stimulus intensity and pain-specific processing [30]. Thus, a null effect of dAI LIFU on TSP does not necessarily argue against dAI engagement or against the insula as an FM target. It may instead indicate that dAI is not the dominant node controlling this lower-level ascending facilitation measure, or that the stimulation dose/timing was insufficient to alter a spinally anchored wind-up process. This is supported by our recent results in a healthy human cohort that found LIFU to only the PI and not the AI affected TSP [22].

#### Why dAI may not strongly affect CPM

CPM is generally used as a proxy for descending antinociceptive control [56,57], but it is not a pure “top-down” test of anterior insula function. CPM involves spinal, brainstem, PAG, cingulate, orbitofrontal, lateral prefrontal, and insular mechanisms [113]. Bogdanov et al. found that behavioral CPM-related hypoalgesia was linked to suppression of posterior insula/SII responses, while early responses in ACC, OFC, and lateral PFC predicted subsequent CPM [113]. This suggests that CPM may depend more directly on prefrontal–cingulate–brainstem descending control and posterior operculo-insular modulation than on dAI alone. Interestingly, our recent work in a healthy cohort found no evidence for changes in CPM for either a PI or AI target [22]. Together, these findings support the idea that dAI is a strong target for FM symptomology broadly, but TSP and CPM may be relatively blunt or anatomically mismatched readouts for detecting acute dAI modulation.

Second, the delivered intracranial pressure in this cohort was substantially lower than that reported in In et al. at comparable anatomical targets [22]. Here, subject-specific acoustic simulations yielded a group-mean peak intracranial pressure of 76.1 ± 25.6 kPa, approximately 42% of the 180.1 ± 84.6 kPa reported by In et al [22]. While this is lower, the minimum effective pressure required for reliable LIFU neuromodulation in humans remains unclear [2], making it difficult to definitively attribute the present null findings to insufficient exposure. However, our prior work provides important context. In et al. delivered higher pressures to the anterior insula using an identical sonication paradigm and likewise observed no effects on either TSP or CPM [22]. In contrast, LIFU to the posterior insula significantly attenuated TSP within the same study. Although differences in delivered pressure between targets may contribute to these effects, these factors were accounted for in their statistical analyses, and the observed dissociation remained. Taken together, these findings suggest that while exposure may influence the magnitude of neuromodulatory effects, the lack of dAI effects on TSP and CPM is more likely explained by target-specific functional anatomy and relevance to FM than by insufficient stimulation intensity alone.

Third, the geometric overlap between the high-pressure region of the beam and the dAI ROI was partial. The dAI ROI received some acoustic pressure in 96.7 ± 7.0% of its voxels, but the high-pressure focal region of the beam, defined as voxels exceeding 50% of the global intracranial peak, covered only 9.3 ± 8.1% of the ROI on average (range 0.0–24.4%), and only 14.1 ± 11.7% of the FWHM beam volume fell within the ROI. Peak pressure at the ROI was 73.8 ± 20.1% of the global intracranial peak, indicating that the highest-pressure point of the beam was not reliably centered on dAI across participants. Importantly, neuronavigation accuracy was < 2mm, so the LIFU trajectory misalignment is not a real-time targeting error but a consequence of how the beam is reshaped after it enters the head due to skull aberration – which is a known limitation of single-element transducers [2]. Despite performing pre-hoc acoustic models to find the best scalp target to place the transducer to maximize overlay of the LIFU beam with the dAI ROI, this target can be challenging due to the complex craniofacial anatomy of the anterior temporal window. In contrast to the relatively thin and uniform squamous temporal bone, anterior placements approach the zygomatic arch, greater wing of the sphenoid, and lateral orbital rim, all of which introduce substantial heterogeneity in skull thickness, curvature, and acoustic impedance. These structures often present as bony protuberances or angled surfaces that can result in refraction, reflection, and phase aberration of the ultrasound beam [2]. As a consequence, the effective acoustic focus may be displaced or attenuated in a subject-specific manner. Because single-element transducers cannot compensate for these skull-induced distortions, targeting of the anterior insula is likely more sensitive to individual anatomical variability, which contributes to variability in target engagement across participants that may help explain the lack of effect.

### Pharmacological Comorbidities in FM Patients

Several features of this cohort distinguish it from the healthy-volunteer samples in which most prior LIFU research has been conducted. Polypharmacy was common: 10 of 13 participants (77%) were taking at least one CNS-active medication at enrollment, including gabapentinoids (4 of 13) and opioid analgesics (1 of 13). Some parallels may be gleaned from other NIBS literature. The safety of TMS at conventional stimulation parameters has been documented in patients taking CNS-active medications [109]. However, medications can modify stimulation-induced cortical plasticity in non-invasive brain stimulation paradigms. For example, for transcranial magnetic and direct current stimulation specifically, medications affecting GABAergic, glutamatergic (NMDA), dopaminergic, serotonergic, cholinergic, and noradrenergic signaling alter plasticity induction in a dose- and receptor-dependent manner [49,50]. Whether these pharmacological principles extend to mechanical neuromodulation has not been systematically investigated. Clinical trials in FM should document, control for, or prospectively analyze CNS-active medication use as a moderator of response, particularly given the high prevalence of gabapentinoids and antidepressants in this population [114,115].

### Limitations

Two limitations warrant acknowledgment. First, the sample was predominantly female (12 of 13) reflecting the established epidemiological skew of FM diagnoses toward women and limiting generalizability to men and to racially and ethnically diverse populations [23]. Future trials should pursue more representative enrollment to characterize whether safety and tolerability profiles differ across demographic groups. Second, the FIQR, which queries fibromyalgia symptom impact over the preceding 7 days, was administered at baseline but not reassessed at a standardized post-LIFU timepoint as there was no follow-up after 72 hours in this study. Its absence limits the ability of this design to detect any sustained effect of LIFU on overall FM symptom burden. Future trials should incorporate serial FIQR assessments at numerous follow-up intervals.

## Conclusions

Here, we establish a feasible, safe, and well-tolerated approach for LIFU in individuals with fibromyalgia. No serious adverse events occurred, no stopping criteria were triggered, participants rated the procedure as comfortable, and blinding integrity was maintained across conditions. Although TSP and CPM did not differ between LIFU and sham, the dorsal anterior insula remains a scientifically motivated target given its role in interoceptive, salience, and integrative aspects of pain processing. Importantly, these findings suggest that neuromodulation of the dAI may be better suited to influence the broader symptom complex of fibromyalgia including pain amplification, multisensory sensitivity, affective distress, fatigue, and interoceptive hypervigilance, rather than evoked pain sensitivity as indexed by TSP and CPM. Future studies should therefore prioritize outcomes that capture these multidimensional features of the disorder to more fully evaluate the therapeutic potential of dAI-targeted LIFU.

## Data Availability

De-identified data supporting the findings of this study may be made available by the corresponding author upon reasonable request, subject to institutional review, applicable data use agreements, and restrictions necessary to protect participant privacy. Individual-level imaging and clinical data are not publicly posted because of human participant privacy considerations.

