## Supplemental Figures and Tables for "Safety and Tolerability of Low Intensity Focused Ultrasound to the Anterior Insula in Patients with Fibromyalgia"

#### SUPPLEMENTAL TABLES

##### Table S1. Acoustic beam profile metrics across participants (N = 13).

| Metric | Mean ± SD | Median | Min | Max |
|---|---|---|---|---|
| Peak intracranial pressure (kPa) | 76.1 ± 25.6 | 78.9 | 27.0 | 123.3 |
| Peak pressure in ROI (kPa) | 56.5 ± 26.9 | 47.3 | 18.0 | 107.2 |
| Mean pressure in ROI (kPa) | 17.9 ± 6.8 | 17.8 | 7.9 | 33.1 |
| Mean pressure in FWHM (kPa) | 48.6 ± 17.1 | 51.0 | 16.8 | 80.8 |
| Peak ROI / peak brain (%) | 73.8 ± 20.1 | 76.1 | 37.4 | 97.6 |
| FWHM beam volume ($mm^3$) | 634 ± 401 | 515 | 192 | 1,650 |
| FWHM beam in ROI ($mm^3$) | 96 ± 76 | 107 | 0 | 229 |
| ROI volume ($mm^3$) | 1,081 ± 119 | 1,095 | 896 | 1,270 |
| ROI covered by FWHM beam (%) | 9.3 ± 8.1 | 8.4 | 0.0 | 24.4 |
| FWHM beam within ROI (%) | 14.1 ± 11.7 | 13.5 | 0.0 | 41.6 |
| ROI receiving any pressure (%) | 96.7 ± 7.0 | 100.0 | 74.7 | 100.0 |

*Note. Peak and mean pressure values are from brain-masked simulations (intracranial voxels only). FWHM = full-width at half-maximum, defined as voxels exceeding 50% of the global intracranial peak pressure. ROI = left dorsal agranular insula (dAI), Brainnetome atlas, 70% probability threshold. Beam dimensions are bounding-box extents of the FWHM mask. "ROI receiving any pressure" quantifies the proportion of ROI voxels where any simulated pressure was present (full beam field, not restricted to FWHM).*

**Table S2. Per-subject neuroradiological review of pre- and post-LIFU structural MRI.**

| Subject | Pre-LIFU (impression) | Post-LIFU (impression) | Change vs. baseline | Stopping criteria triggered |
|---|---|---|---|---|
| S01 | Essentially unremarkable noncontrast MRI of the brain | Essentially unremarkable noncontrast MRI of the brain | Stable; isolated focus of perivascular gliosis (no significance) unchanged | No |
| S02 | Essentially unremarkable noncontrast brain protocol MRI | Essentially unremarkable noncontrast brain protocol MRI | Stable; very minimal bifrontal subcortical leukoaraiosis (typical for age) unchanged | No |
| S03 | Benign-appearing prepontine cystic lesion (ecchordosis physaliphora favored); minimal white matter changes | Unchanged: benign-appearing prepontine cystic lesion; minimal white matter changes | Unchanged | No |
| S04 | Essentially unremarkable noncontrast brain protocol MRI | — | — | — |
| S05 | Slight senescent change (typical for age); cervical spondylosis at study periphery | Similar: slight senescent change (typical for age); cervical spondylosis at study periphery | Similar; essentially unchanged | No |
| S06 | Senescent change (typical for age); otherwise essentially unremarkable | Essentially unchanged: senescent change (typical for age); otherwise essentially unremarkable | Essentially unchanged | No |
| S07 | Essentially unremarkable noncontrast brain protocol MRI | Minimal senescent change (typical or less than typical for age); left maxillary mucosal disease; cervical spondylosis | Minimal leukoaraiosis better delineated on post-LIFU than on baseline; attributed to technical factors by neuroradiologist. No new CNS findings. | No |
| S08 | Essentially unremarkable noncontrast brain protocol MRI (0.6 cm pineal cyst, common normal variant) | Mild paranasal sinus mucosal disease; unchanged pineal cyst; otherwise unremarkable | Pineal cyst (common normal variant) unchanged; new paranasal sinus mucosal thickening (non-CNS) | No |
| S09 | Essentially unremarkable noncontrast brain protocol MRI (subcentimeter right choroid plexus xanthogranuloma, no significance) | — | — | — |

| Subject | Pre-LIFU (impression) | Post-LIFU (impression) | Change vs. baseline | Stopping criteria triggered |
|---|---|---|---|---|
| S10 | — | — | — | — |
| S11 | Essentially unremarkable noncontrast brain protocol MRI (slightly prominent perivascular space, no significance) | Essentially unremarkable noncontrast brain protocol MRI | Stable; prominent perivascular space (no significance) unchanged | No |
| S12 | Normal noncontrast MRI of the brain | Normal noncontrast MRI of the brain | Unchanged; normal at both timepoints | No |
| S13 | Senescent change (typical for age); cervical spondylosis at study periphery | Essentially unchanged: senescent change (typical for age); cervical spondylosis | Essentially unchanged | No |

*Em-dashes (—) indicate cells for which neuroradiology reads are not yet available.*

### SUPPLEMENTAL FIGURES

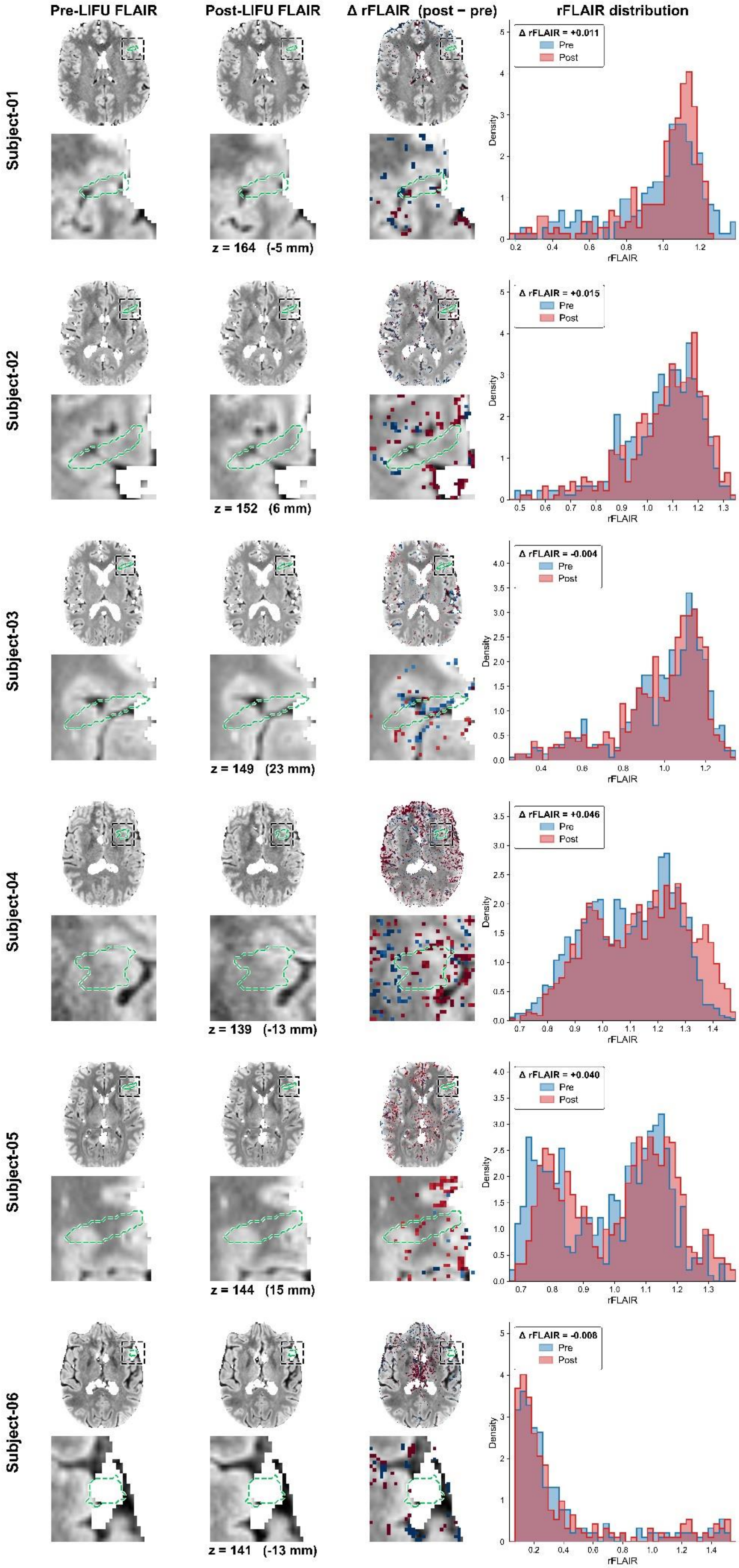

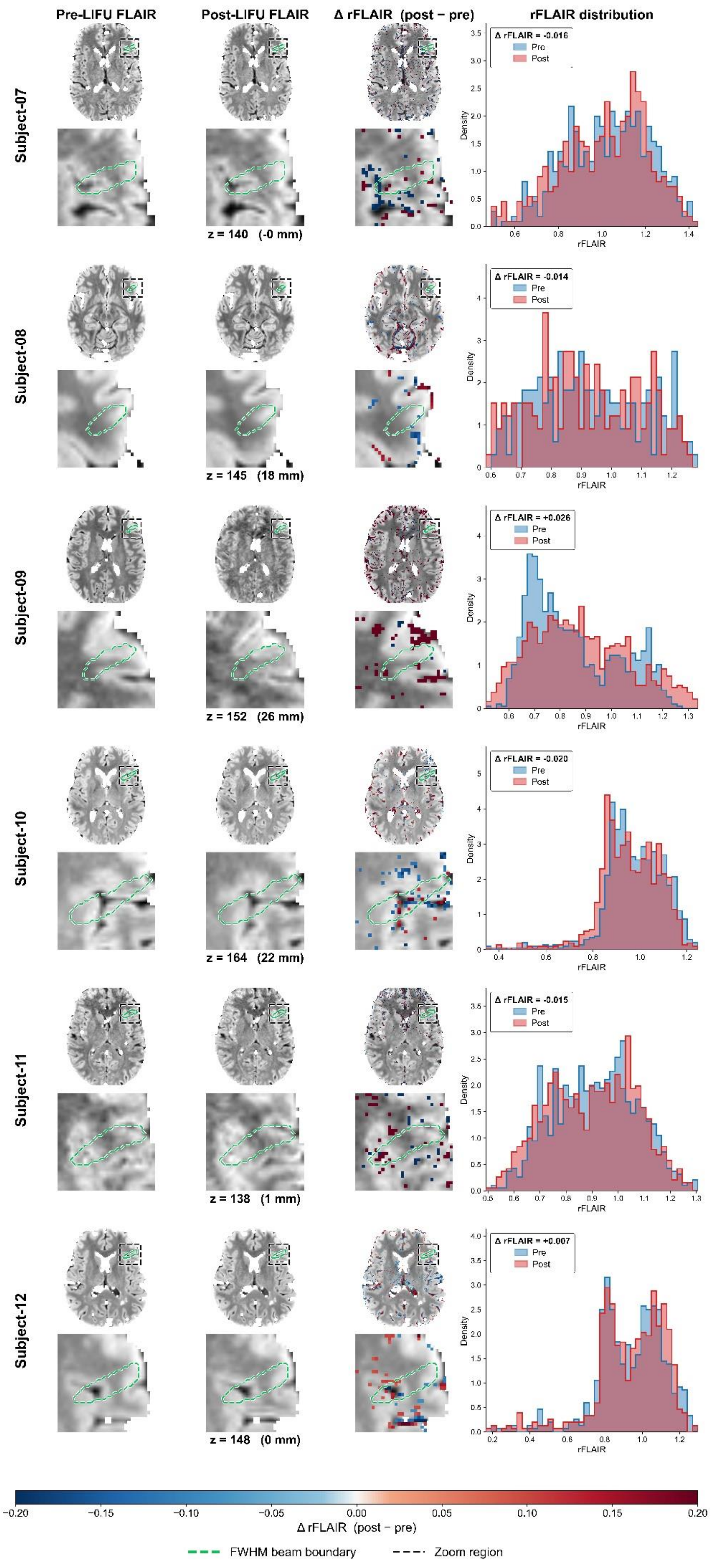

Pre-LIFU FLAIR
Post-LIFU FLAIR
Δ rFLAIR (post − pre)
rFLAIR distribution
Subject-07
Δ rFLAIR = -0.016
Pre
Post
Density
rFLAIR
z = 140 (-0 mm)
Subject-08
Δ rFLAIR = -0.014
z = 145 (18 mm)
Subject-09
Δ rFLAIR = +0.026
z = 152 (26 mm)
Subject-10
Δ rFLAIR = -0.020
z = 164 (22 mm)
Subject-11
Δ rFLAIR = -0.015
z = 138 (1 mm)
Subject-12
Δ rFLAIR = +0.007
z = 148 (0 mm)
−0.20
−0.15
−0.10
−0.05
0.00
0.05
0.10
0.15
0.20
Δ rFLAIR (post − pre)
FWHM beam boundary
Zoom region

**Supplemental Figure 1. Per-subject voxel-wise rFLAIR analysis across the cohort.** Each row corresponds to one participant and presents, from left to right: the pre-LIFU FLAIR, the post-LIFU FLAIR, the voxel-wise ΔrFLAIR (post − pre) overlaid on the pre-LIFU FLAIR image, and the within-FWHM beam rFLAIR distribution (overlaid normalized histograms; pre, blue; post, red; per-subject mean ΔrFLAIR shown in the top-left of each panel). Within each row, the first three columns show an axial full-brain slice (top) and a zoomed inset at the LIFU target (bottom) at the slice z-coordinate indicated below the post-LIFU panel. The dashed magenta-pink contour outlines the FWHM beam boundary, and the dashed rectangle in the full-brain view marks the inset region. The horizontal colorbar at the bottom of the figure spans the ΔrFLAIR scale used in column 3.
*Note. For layout clarity, this figure shows 12 of the 13 cohort participants; one participant (Subject S07) is omitted from the display but is included in all statistical analyses reported in the main text, with their values reported in the S07 row of Table 2, and visualization reported in the Figure 2.*

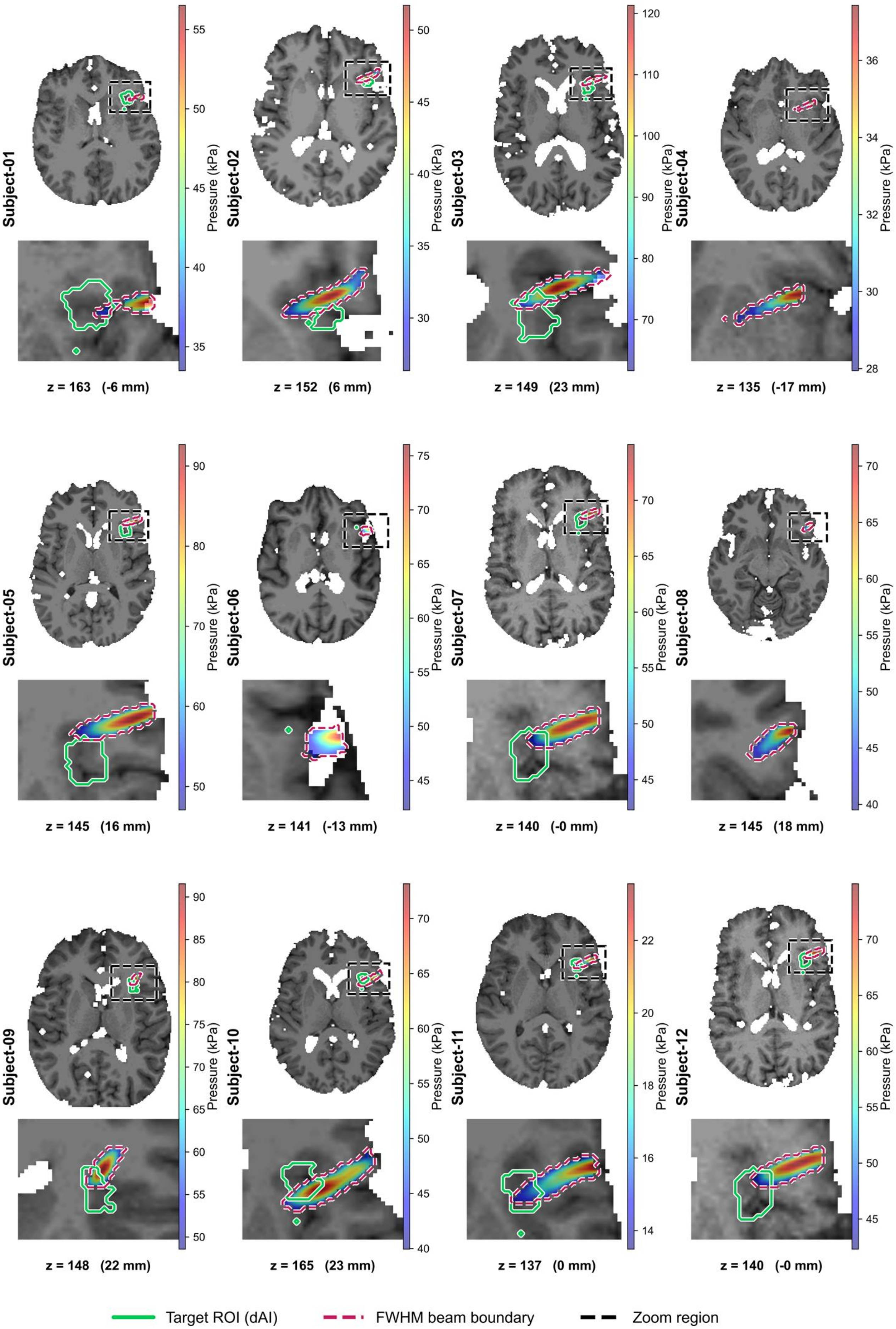
Subject-01
Pressure (kPa)
z = 163 (-6 mm)
Subject-02
Pressure (kPa)
z = 152 (6 mm)
Subject-03
Pressure (kPa)
z = 149 (23 mm)
Subject-04
Pressure (kPa)
z = 135 (-17 mm)
Subject-05
Pressure (kPa)
z = 145 (16 mm)
Subject-06
Pressure (kPa)
z = 141 (-13 mm)
Subject-07
Pressure (kPa)
z = 140 (-0 mm)
Subject-08
Pressure (kPa)
z = 145 (18 mm)
Subject-09
Pressure (kPa)
z = 148 (22 mm)
Subject-10
Pressure (kPa)
z = 165 (23 mm)
Subject-11
Pressure (kPa)
z = 137 (0 mm)
Subject-12
Pressure (kPa)
z = 140 (-0 mm)
Target ROI (dAI)
FWHM beam boundary
Zoom region

2.

**Supplementary Fig. 2. Per-participant acoustic simulation across the cohort**. For each participant, the upper panel shows the simulated intracranial acoustic pressure field (jet colormap) overlaid on the participant's T1-weighted image, with the left dorsal agranular insula (lh-dAI) target outlined in solid green and the full-width at half-maximum (FWHM) beam boundary (voxels ≥ 50% of maximum intracranial pressure) in dashed magenta-pink. The lower panel shows a zoomed inset at the target (location marked by the dashed rectangle in the upper image). The pressure colorbar to the right of each panel is scaled to that participant's individual peak intracranial pressure (kPa); the slice z-coordinate is indicated beneath each panel.

*Note. For layout clarity, this figure shows 12 of the 13 cohort participants; the omitted participant (Subject S07 in Supplementary Table 1) is included in all reported analyses, with their values reported in the S07 row of Supplementary Table 1.*
